# Causal inference using observational intensive care unit data: a systematic review and recommendations for future practice

**DOI:** 10.1101/2022.10.29.22281684

**Authors:** J. M. Smit, J. H. Krijthe, J. van Bommel, J.A. Labrecque, M. Komorowski, D.A.M.P.J. Gommers, M. J. T. Reinders, M.E. van Genderen

## Abstract

**Aim:** To review and appraise the quality of studies that present models for causal inference of time-varying treatment effects in the adult intensive care unit (ICU) and give recommendations to improve future research practice.

**Methods:** We searched Embase, MEDLINE ALL, Web of Science Core Collection, Google Scholar, medRxiv, and bioRxiv up to March 2, 2022. Studies that present models for causal inference that deal with time-varying treatments in adult ICU patients were included. From the included studies, data was extracted about the study setting and applied methodology. Quality of reporting (QOR) of target trial components and causal assumptions (ie, conditional exchangeability, positivity and consistency) were assessed.

**Results:** 1,714 titles were screened and 60 studies were included, of which 36 (60%) were published in the last 5 years. G methods were the most commonly used (n=40/60, 67%), further divided into inverse-probability-of-treatment weighting (n=36/40, 90%) and the parametric G formula (n=4/40, 10%). The remaining studies (n=20/60, 33%) used reinforcement learning methods. Overall, most studies (n=36/60, 60%) considered static treatment regimes. Only ten (17%) studies fully reported all five target trial components (ie, eligibility criteria, treatment strategies, follow-up period, outcome and analysis plan). The ‘treatment strategies’ and ‘analysis plan’ components were not (fully) reported in 38% and 48% of the studies, respectively. The ‘causal assumptions’ (ie, conditional exchangeability, positivity and consistency) remained unmentioned in 35%, 68% and 88% of the studies, respectively. All three causal assumptions were mentioned (or a check for potential violations was reported) in only six (10%) studies. Sixteen studies (27%) estimated the treatment effect both by adjusting for baseline confounding and by adjusting for baseline and treatment-affected time-varying confounding, which often led to substantial changes in treatment effect estimates.

**Conclusions:** Studies that present models for causal inference in the ICU were found to have incomplete or missing reporting of target trial components and causal assumptions. To achieve actionable artificial intelligence in the ICU, we advocate careful consideration of the causal question of interest, the use of target trial emulation, usage of appropriate causal inference methods and acknowledgement (and ideally examination of potential violations) of the causal assumptions.

**Systematic review registration:** PROSPERO (CRD42022324014)

## Introduction

Many treatment choices in the intensive care unit (ICU) are made quickly, based on patient characteristics that are changing and monitored in real-time. Given this dynamic and data-rich environment, the ICU is pre-eminently a place where artificial intelligence (AI) holds the promise to aid clinical decision making.^1–3^ So far, however, most AI models developed for the ICU remain within the prototyping phase.^4,5^ One explanation for this may be that most models in the ICU are built for the task of prediction, ie, mapping input data to (future) patient outcomes.^6^ However, even a very accurate prediction of, for instance, sepsis,^7^ does not tell a physician what to do in order to treat or prevent it. In other words, prediction models are seldom actionable. For AI that assists clinicians in what to do, ie, ‘actionable AI’, models need to take into account cause and effect.

Causal inference (CI) represents the task of estimating causal effects by comparing patient outcomes under multiple counterfactual treatments.^6,8^ The most widely used method for CI is a randomized controlled trial (RCT). Through randomization (coupled with full compliance), the difference in outcome between treatment groups can be interpreted as a causal treatment effect. Because carrying out RCTs may be infeasible due to cost, time, and ethical constraints, observational studies are sometimes the only alternative. CI using observational data can be seen as an attempt to emulate the RCT that would have answered the question of interest (ie, the ‘target trial’).^9^ With such an approach, however, treatment is not assigned randomly and extra adjustment for confounding is required. In the simple situation of a time-fixed (or ‘point’) treatment (figure 1, panel 1),^10,11^ this can be achieved by ‘standard methods’ like regression or propensity-score (PS) analyses.^12^ However, ICU physicians are typically confronted with treatment decisions which occur at multiple time-points, ie, time-varying treatments (figure 1, panel 1).^10,11^ Estimating the effect of time-varying treatments using observational data is often complicated by treatment-confounder feedback,^13^ leading to ‘treatment-affected time-varying confounding’ (TTC, panel 1)^11,14,15^. Usage of standard methods in the presence of TTC leads to bias.^16,17^ Inverse-probability-of-treatment weighting (IPTW), the parametric G formula and G estimation (collectively known as ‘G methods’, panel 1) were introduced by Robins^18^ to estimate causal effects in the presence of TTC, making them well-suited for CI in the ICU. Time-varying treatments can be further subdivided into static (STRs) and dynamic treatment regimes (DTRs, figure 1, panel 1). The latter type is most common in the ICU, as treatment choices are typically dynamically re-evaluated based on the evolving patient state. For example, rather than deciding upon admission to administer vasopressors daily, an ICU physician reconsiders giving this treatment throughout the ICU stay based on the patient’s response. Hence, the practical question of interest often requires a comparison of DTRs. Reinforcement learning (RL)^19^ is another class of methods which, like G methods, can be used to estimate optimal DTRs and have been increasingly applied to ICU data.^20^ Partly due to the different language used to describe similar concepts (table S1), studies applying G methods and RL may appear as completely separate disciplines. However, they show great similarities and can be used to build actionable AI models.

**Figure 1:**
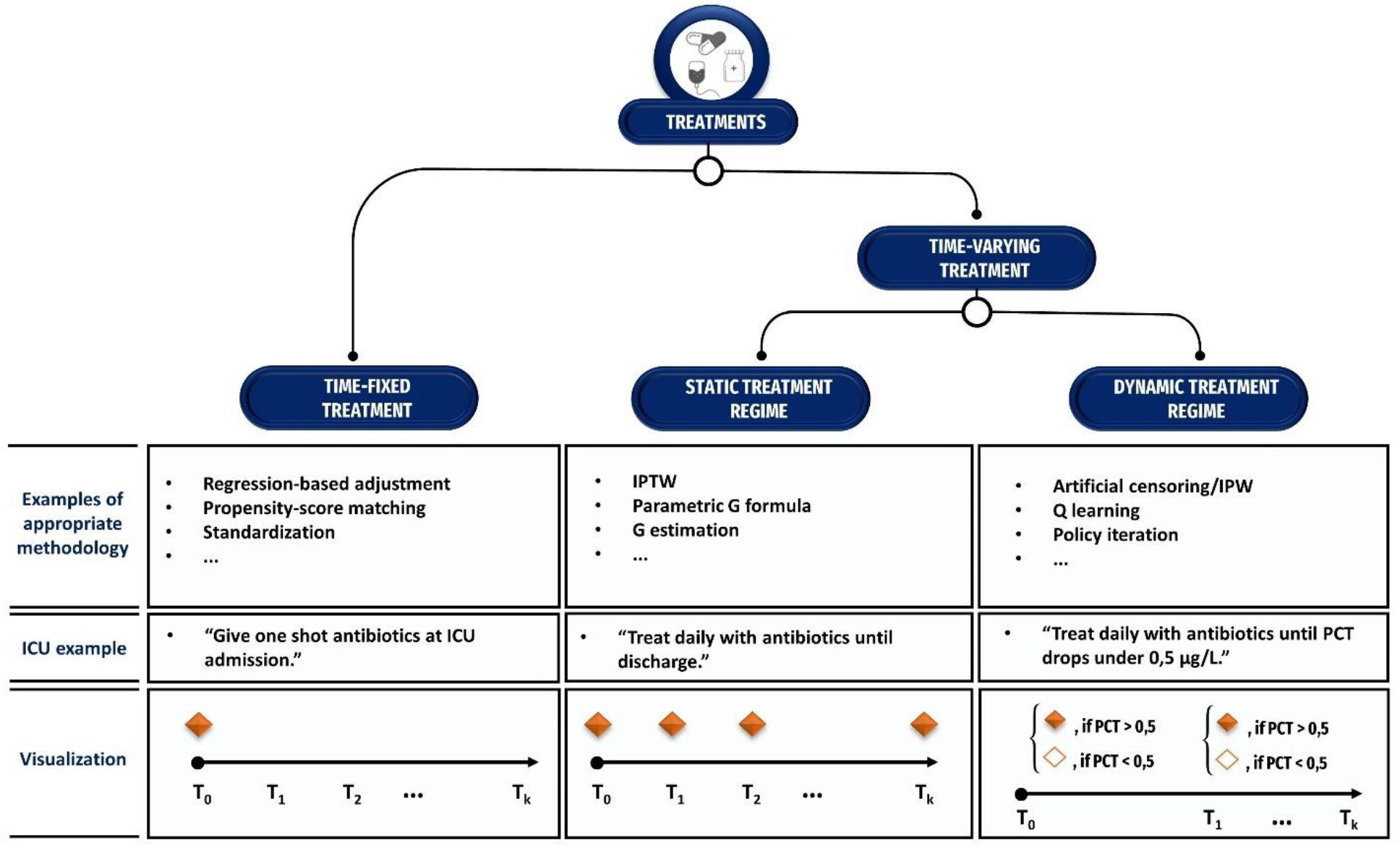
Taxonomy of treatment types. Treatments can be time-fixed or time-varying, and time-varying treatments can be subdivided into static and dynamic treatment strategies. Appropriate methodology to estimate causal effects of treatment based on observational data depends on the treatment type. ICU=intensive care unit, IPTW=inverse-probability-of-treatment weighting, IPW=inverse-probability weighting, CRP=C-reactive protein.

To move towards actionable AI in the ICU, our review provides an overview of CI studies concerning time-varying treatments in the ICU, discusses quality of reporting and gives recommendations to improve future research practice.

### Panel 1: Glossary

#### Time-fixed treatment

a treatment that only occurs at the start of follow-up (eg, one shot antibiotics at ICU admission), or does not change over time (eg, genotype).

#### Time-varying treatment

any treatment that is not time-fixed. Time-varying treatments can be sub-divided in static and dynamic treatment regimes.

#### Static treatment regime (STR)

a treatment regime that is not tailored to evolving patient characteristics (eg, ‘treat patient with daily antibiotics during ICU admission’).

#### Dynamic treatment regime (DTR)

a treatment regime where the treatment decisions depend on changing patient characteristics and/or treatment history (eg, ‘treat patient with antibiotics until procalcitonin drops below 0.5 μg/L’).

#### Treatment-affected time-varying confounding (TTC)

time-varying confounding in which one or more time-varying confounders are affected by previous treatment.

#### G methods

a class of methods proposed to appropriately adjust for TTC in the estimation of time-varying treatment effects, including inverse-probability-of-treatment weighting (IPTW), the parametric G formula, and G estimation.

#### Reinforcement learning (RL)

a class of methods that deals with the problem of sequential decision making which returns an optimized treatment regime, including (among others) Q-learning and policy iteration.

#### Off-policy evaluation (OPE)

the task of estimating the value of an (optimized) treatment regime (or ‘policy’) using data from patients who received treatments not conform to this regime (eg, observational data). OPE methods fall into two main categories: importance-sampling and model-based methods (doubly robust methods borrow ideas from both importance-sampling and model-based methods).

### Causal assumptions

#### Conditional exchangeability

Exchangeability means that the risk of an outcome (eg, mortality) in the untreated group (eg, those who did not receive antibiotics) would have been the same as the risk in the treated group (eg, those who received antibiotics), had the patients in the untreated group received treatment. In observational data, exchangeability generally does not hold due to confounding and/or selection bias, and, therefore, CI requires the assumption that all confounders are measured and adjusted for to achieve exchangeability *conditional* on the measured confounders.

##### Positivity

One can only estimate the causal effect of a treatment by comparing data of treated and untreated patients. Therefore, in all subgroups (or ‘strata’) defined by specific combinations of the confounder values, there must be treated and untreated patients. In other words, treatment should occur with some *positive* probability in all confounder strata (ie, the *positivity* assumption).

##### Consistency

Consistency assumes that the outcome for a given treatment will be the same, irrespective of the way treatments are ‘assigned’. This is often plausible for medical treatments, but less obvious for treatments that are modifiable by a variety of means, such as body-mass index which could be caused by diet or a metabolic disorder.

## Methods

This systematic review was conducted in accordance with the Preferred Reporting Items for Systematic Reviews and Meta-Analysis (PRISMA) guidelines,^21^ and the protocol was registered in the online PROSPERO database (CRD42022324014).^22^

### Search strategy

Candidate articles were identified through a comprehensive search in Embase, MEDLINE ALL, Web of Science Core Collection, Google Scholar, medRxiv, and bioRxiv up to 2 March 2022, with no start date. We developed a search strategy that could be modified for each database (appendix A). Search terms included relevant controlled vocabulary terms and free text variations for CI, G methods, or common RL methods, combined with ICU related terms.

### Eligibility criteria

We included any primary research article, conference proceedings or pre-print papers that present models for the task of CI concerning time-varying treatments in adult (≥ 17 years of age) patients admitted to the ICU. Articles were not eligible if data from an RCT was used (unless the treatment of interest was not the randomized treatment), it focused on the introduction of new methodology, or was an abstract-only, review, opinion, or survey. Duplicates and articles not written in English were also excluded.

### Study selection

We used a two-stage approach for screening: first by title and abstract and then by full article text. One reviewer (JS) screened the titles and abstracts. Full text articles were then screened and selected. Studies for which uncertainty remained for eligibility were independently screened in full-text by two other reviewers (JK and MvG), and conflicts were resolved by discussion between the three reviewers. For both title-abstract and full text screening, reasons for exclusion were recorded.

### Data extraction

Data was extracted by using a standardised data extraction form. Uncertainties were resolved by discussion between three reviewers (JS, JK and JL). The items for extraction were based on the STROBE (STrengthening the Reporting of OBservational studies in Epidemiology) checklist,^23^ supplemented by method-specific items. We extracted the following items from all included studies: details of study variables (ie, studied treatment and primary outcome), the number of included ICUs, usage of open-source database(s), number of participants included, studied treatment type (figure 1), and used CI method. In addition, we extracted the following method-specific items: the usage of methods to reduce extreme weights (ie, weight stabilization^24^ and truncation^25^) for studies using IPTW and the off-policy evaluation^26^ (OPE, panel 1) method used for studies using RL. Finally, if a study estimated the treatment effect both by adjusting for baseline confounding and by adjusting for baseline confounding and TTC, we also collected these different estimates.

### Quality of reporting

To assess the quality of reporting (QOR) of the included studies, we judged the reporting of the components of the target trial framework^9^ and the causal assumptions (panel 1).

#### Target trial components

The ‘target trial framework’, introduced by Hernan and Robins^9^, consists of seven main components. We judged the reporting of five of these: eligibility criteria, treatment strategies, follow-up period, outcome and analysis plan (table 1). We scored the analysis plan component as ‘reported’ if one could reproduce the modelling given the required data. For studies using RL, we judged the ‘treatment strategies’ and ‘outcome’ components as ‘reported’ if the definitions of the used action space and reward were reported, respectively. We split the follow-up period component into three subcomponents: time-zero (or ‘index date’), end of follow-up, and time resolution (ie, the time steps in which the treatment level is considered the same).^27^ We split the analysis plan component into specific subcomponents depending on the CI method used (table S2). We scored the target trial components that are split in subcomponents as ‘reported, ‘partially reported’ and ‘not reported’ if all, some, or none of the subcomponents were reported, respectively. Because (outside RCTs) ICU physicians are typically aware of the treatment patients receive, one cannot emulate target trials with blind assignment. Therefore, we did not consider the ‘assignment procedures’ component. Also, we did not consider the ‘causal contrast of interest’ component because an intention-to-treat analysis based on observational data is rarely possible.^9^

**Table 1:** Components of the target trial framework included in the QOR assessment, including examples for the ICU setting. *The analysis plan component is subdivided in a specific set of subcomponents depending on the modelling strategy used, these are summarized in table S2. **This description would not be considered as sufficient reporting of the analysis plan component, but simply serves as an example.

| Component | Subcomponent | Included for QOR assessment | ICU example |
| --- | --- | --- | --- |
| Eligibility criteria | - | ✓ | Individuals aged 65 years or older admitted to the ICU meeting Sepsis-3 criteria upon admission. |
| Treatment strategies | - | ✓ | 1. administration of 50-75 mL/kg of intravenous fluid boluses during the first several hours of treatment (ie, liberal fluid therapy<sup>80</sup>)<br>2. Intravenous fluid boluses of 250–500 mL during ICU stay in the case of severe hypoperfusion (ie, restrictive fluid therapy<sup>81</sup>) |
| Assignment procedures | - |  | Unblinded random assignment to one of the treatment strategies. |
| Follow-up period | Start of follow-up | ✓ | Time of ICU admission. |
|  | End of follow-up | ✓ | Death, ICU discharge, or loss to follow-up, whichever occurs first. |
|  | Time resolution | ✓ | Hourly weights were applied to adjust for the impact of time-varying confounders on the hourly risk of adhering to one of the two treatment strategies. |
| Outcome | - | ✓ | All- cause mortality. |
| Causal contrasts of interest | - |  | Per-protocol effect. |
| Analysis plan | * | ✓ | Per-protocol effect will be estimated adjusting for pre- and postbaseline confounders by a marginal structural model using IPTW.** |

#### Causal assumptions

The task of CI relies on strong assumptions, including conditional exchangeability, positivity and consistency (hereafter referred to as ‘causal assumptions’, panel 1).^8^ Violations of these assumptions lead to biased estimates and therefore, acknowledgement is important and ideally, potential violations are examined. We scored each study using three levels of increasingly good reporting quality: (1) assumption not mentioned, (2) assumption mentioned, and (3) attempt to check for potential violations of the assumption reported. For the conditional exchangeability assumption, we distinguished two types of attempts to check for potential violations: ‘indirect approaches’^9^ and ‘bias analyses’.^28^ For the positivity assumption, we considered the examination of the distribution of the estimated (stabilized) inverse-probability-of-treatment (IPT) weights as an attempt to check for potential violations.^29^ Approaches to check potential violations of consistency do not exist and therefore, mentioning the consistency assumption (level 2) was considered as the highest level of reporting quality.

### Evidence synthesis

We tabulated extracted study items for each study individually and grouped by CI method used. QOR results concerning the target trial components and the causal assumptions are summarised as percentages using bar charts, and QOR results for each study individually were tabulated. For the reporting of the target trial components, we made separate tables for each group of studies that used a specific CI method. The collected treatment effect estimates reported by studies that estimated the treatment effect both by adjusting for baseline confounding and by adjusting for baseline and TTC, were plotted as point estimates and corresponding 95% confidence intervals.

## Results

Our search identified 1,714 unique articles, of which 1,605 were excluded based on title and abstract screening. We screened 109 full texts, 60 of which met the eligibility criteria and were included in the review (figure 2). The articles were published between 2005 and 2021 in 36 different journals and conference proceedings, with a steadily growing number of articles per year starting around 2010 (figure S1). A reference list of all included studies and the list with collected items per study can be found in appendix B and table S3, respectively. Most studies applied G methods (n=40, 67%), of which 36 (60%) used IPTW and four (7%) the parametric G formula. Twenty (33%) studies used RL methods (table 2). The three most frequently studied treatment categories were nosocomial infections (n=8, 13%), anti-inflammatory drugs (n=6, 10%) and sedatives and analgesics (n=6, 10%). Most studies (n=32, 53%) considered mortality (at varying follow-up times) as the primary outcome. Thirty-one studies (52%) included data from at least two different ICUs. Studies that used RL generally included more patients than studies that used G methods, with a median of 7,513 (IQR 5,252 to 18,340) versus 1,451 patients (IQR 421 to 2,914) and relied more often on open-source ICU databases (75% vs 15%). In total, 21 (35%) of the studies used one or more open-source ICU database, among which the Medical Information Mart for Intensive Care (MIMIC)-III database^30^ was the most frequently used (n=19, 32%). In contrast to RL studies (which inherently deal with DTRs), only three^31–33^ of the 40 studies (8%) that used G methods considered DTRs.

**Table 2:** Characteristics of the reviewed studies, grouped by used causal inference method. *For the number of included ICUs and study size, the studies that used simulated patient data (n=4) are not taken into account and therefore, the number of studies add up to 16 and 56, respectively.

|  | **IPTW (N=36)**<br>**No (%)** | **Parametric G**<br>**formula (N=4)**<br>**No (%)** | **RL (N=20\*)**<br>**No (%)** | **All (N=60\*)**<br>**No (%)** |
| --- | --- | --- | --- | --- |
| **Studied treatment (categorized)** |  |  |  |  |
| Nosocomial infections | 8 (22) | 0 (0) | 0 (0) | 8 (13) |
| Anti-inflammatory drugs | 5 (14) | 0 (0) | 1 (5) | 6 (10) |
| Sedatives & analgesics | 1 (3) | 0 (0) | 5 (25) | 6 (10) |
| Vasopressors & intra-venous fluids | 0 (0) | 0 (0) | 5 (25) | 5 (8) |
| Antimicrobials | 4 (11) | 0 (0) | 0 (0) | 4 (7) |
| Mechanical ventilation | 0 (0) | 1 (25) | 2 (10) | 3 (5) |
| Anticoagulants | 1 (3) | 0 (0) | 2 (10) | 3 (5) |
| Diuretics | 3 (8) | 0 (0) | 0 (0) | 3 (5) |
| Renal replacement therapy | 3 (8) | 0 (0) | 0 (0) | 3 (5) |
| Other | 11 (31) | 3 (75) | 5 (25) | 19 (32) |
| **Primary outcome (categorized)** |  |  |  |  |
| Mortality | 25 (69) | 1 (25) | 6 (30) | 32 (53) |
| Combined | 0 (0) | 0 (0) | 7 (35) | 7 (12) |
| Maintenance of clinical target value | 0 (0) | 0 (0) | 5 (25) | 5 (8) |
| Nosocomial infections | 1 (3) | 2 (50) | 0 (0) | 3 (5) |
| Need for mechanical ventilation | 2 (6) | 0 (0) | 0 (0) | 2 (3) |
| Other | 8 (22) | 1 (25) | 2 (10) | 11 (18) |
| **Number of included ICUs** |  |  |  |  |
| 1 | 14 (39) | 2 (50) | 13 (81) | 29 (52) |
| 2-4 | 5 (14) | 0 (0) | 1 (6) | 6 (11) |
| 5-10 | 4 (11) | 0 (0) | 0 (0) | 4 (7) |
| 11-20 | 6 (17) | 1 (25) | 0 (0) | 7 (12) |
| 21-100 | 2 (6) | 1 (25) | 0 (0) | 3 (5) |
| >100 | 5 (14) | 0 (0) | 2 (12) | 7 (12) |
| **Utilised open source databases** |  |  |  |  |
| MIMIC-II | 0 (0) | 0 (0) | 1 (5) | 1 (2) |
| MIMIC-III | 6 (17) | 0 (0) | 13 (65) | 19 (32) |
| MIMIC-IV | 0 (0) | 0 (0) | 1 (5) | 1 (2) |
| eICU | 0 (0) | 0 (0) | 2 (10) | 2 (3) |
| AmsterdamUMCdb | 0 (0) | 0 (0) | 1 (5) | 1 (2) |
| **Study size (n patients)** |  |  |  |  |
| 0-100 | 2 (6) | 0 (0) | 0 (0) | 2 (4) |
| 100-500 | 8 (22) | 1 (25) | 0 (0) | 9 (16) |
| 501-1000 | 6 (17) | 0 (0) | 0 (0) | 6 (11) |
| 1001-5000 | 16 (44) | 2 (50) | 4 (25) | 22 (39) |
| >5000 | 4 (11) | 1 (25) | 12 (75) | 17 (30) |
| **Type of treatment regime** |  |  |  |  |
| Static | 34 (94) | 3 (75) | 0 (0) | 37 (62) |
| Dynamic | 2 (6) | 1 (25) | 20 (100) | 23 (38) |

**Figure 2:**
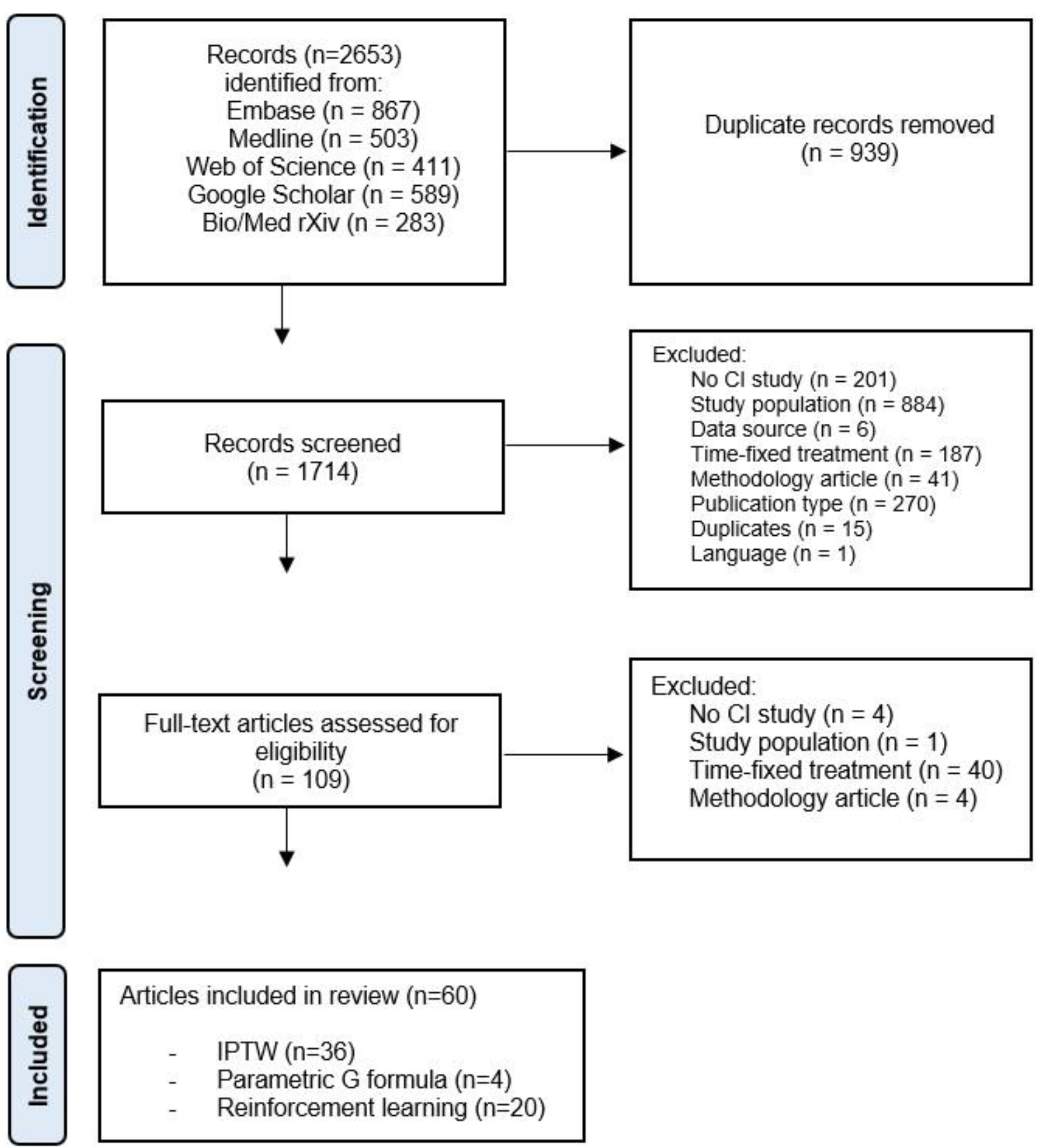
Flowchart of study selection.

### Method-specific items

Among the studies that used IPTW (n=36), 17 applied stabilized weights, one applied weight truncation, and eight studies applied both weight stabilization and truncation. Among studies that applied RL on real (ie, not simulated) patient data (n=16), seven studies used an importance-sampling based^34^, model-based^35,36^, a doubly robust OPE method^37^, or a combination of these. Eight studies used the so-called ‘U-curve method’^38^ (panel 1) and for six of these, this was the only reported OPE method. In three studies, the OPE method was not reported (figure S2).

### Quality of reporting

#### Target trial components

The ‘eligibility criteria’ and ‘outcome’ components were reported in 58 (97%) and 59 (98%) of the studies, respectively (figure 3a). We scored the ‘treatment strategies’, ‘follow-up period’ and ‘analysis plan’ components as partially or not reported in respectively 23 (38%), 16 (27%) and 29 (48%) of the studies. All five target trial components were fully reported in only ten (17%) studies.^31,39–47^ The reporting of the target trial components grouped by used CI method are summarized in figures S3-5 and tabulated for each individual study in tables S4-S6.

**Figure 3:**
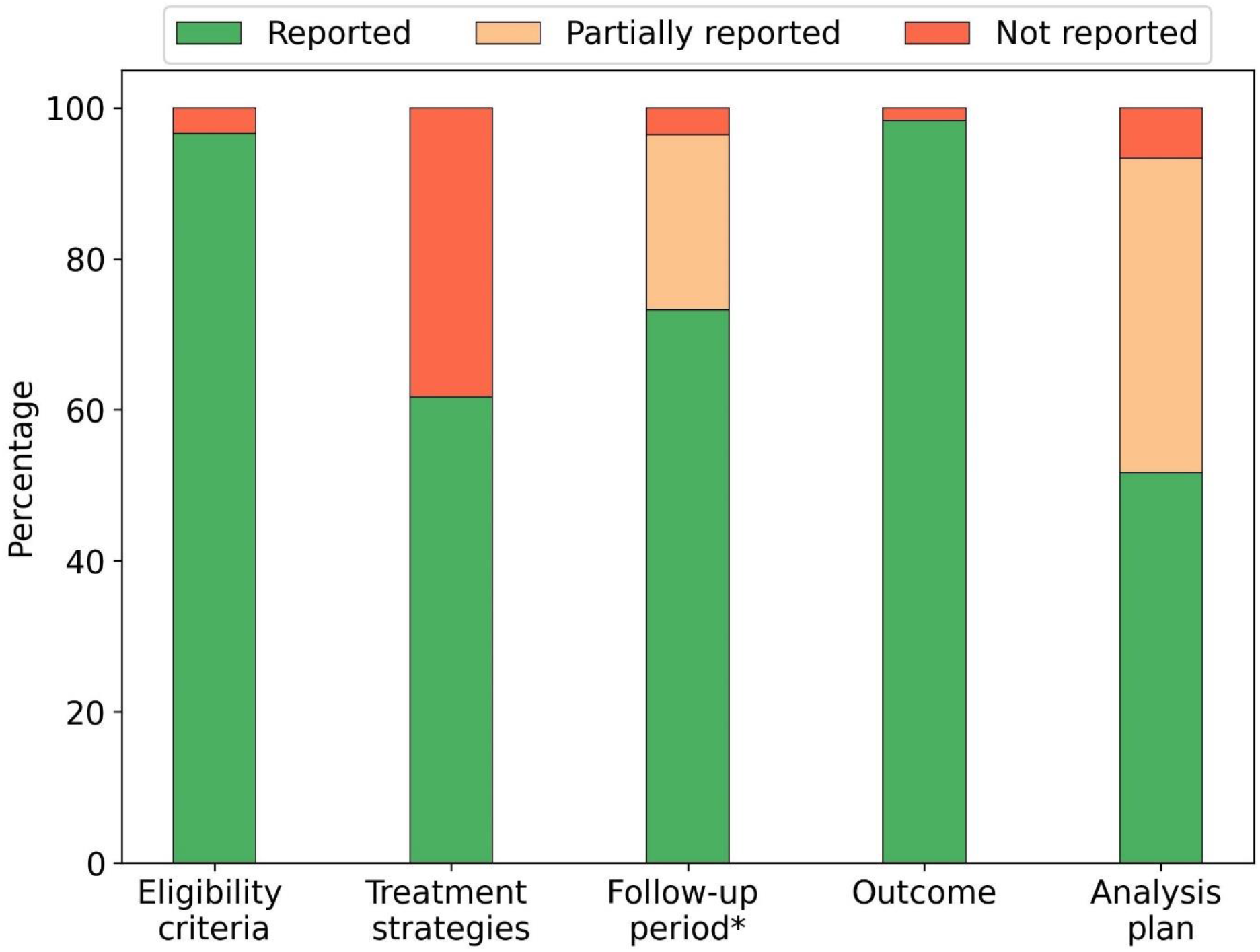
QOR summary plots: reporting of (a) the target trial components and (b) causal assumptions. Figure 3a: Reporting of target trial components. *For the follow-up component, the studies that used simulated patient data (n=4) are not taken into account.

#### Causal assumptions

The conditional exchangeability assumption remained unmentioned in 21 (35%), was mentioned in 25 (42%), and an attempt to check for potential violations was reported in 14 studies (23%, figure 3b). Among the studies that reported a check for potential violations, four studies^48–51^ performed a bias analysis. The positivity assumption remained unmentioned in 41 (68%), was mentioned in three (5%), and a check for potential violations was reported in 16 (27%) of the studies. The consistency assumption was mentioned in seven (12%) of the studies. All three assumptions were mentioned (or a check for potential violations was reported) in only six (10%) studies.^31,33,42,43,51,52^ The reporting of assumptions grouped by CI method used are summarised in figures S6-S8 and individual results for all studies are tabulated in table S7. In general, the causal assumptions remained unmentioned more often in studies that applied RL, compared to those which applied G methods (figures S6-8). All studies that reported a check for potential violations of the conditional exchangeability assumption also mentioned this assumption, whereas for the positivity assumption, seven out of 16 studies that reported a check for potential violations did not explicitly mention positivity (table S7).

**Figure 3b:**
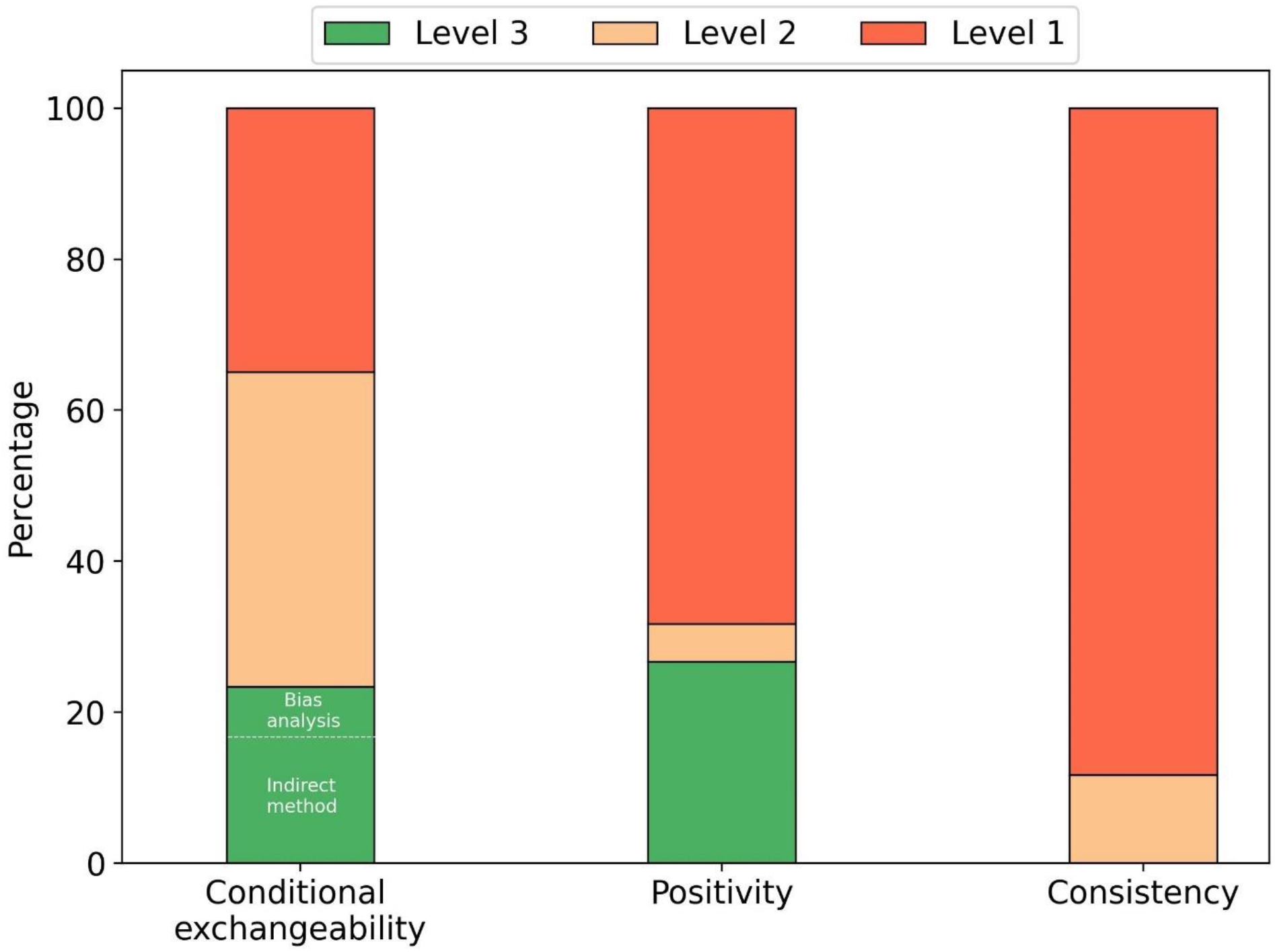
Reporting of causal assumptions. Level 1=assumption not mentioned, level 2=assumption mentioned, level 3=attempt to check for potential violations of the assumption reported.

### Adjusting for TTC

Eighteen studies (30%) estimated the treatment effect by adjusting for baseline confounding and by adjusting for baseline confounding and TTC. For most of these studies, the point estimates of the treatment effects varied substantially after adjusting for both baseline and TTC, moving the effect estimate towards or away from the null hypothesis, or even leading to opposite effect estimates (figure 4).

**Figure 4:**
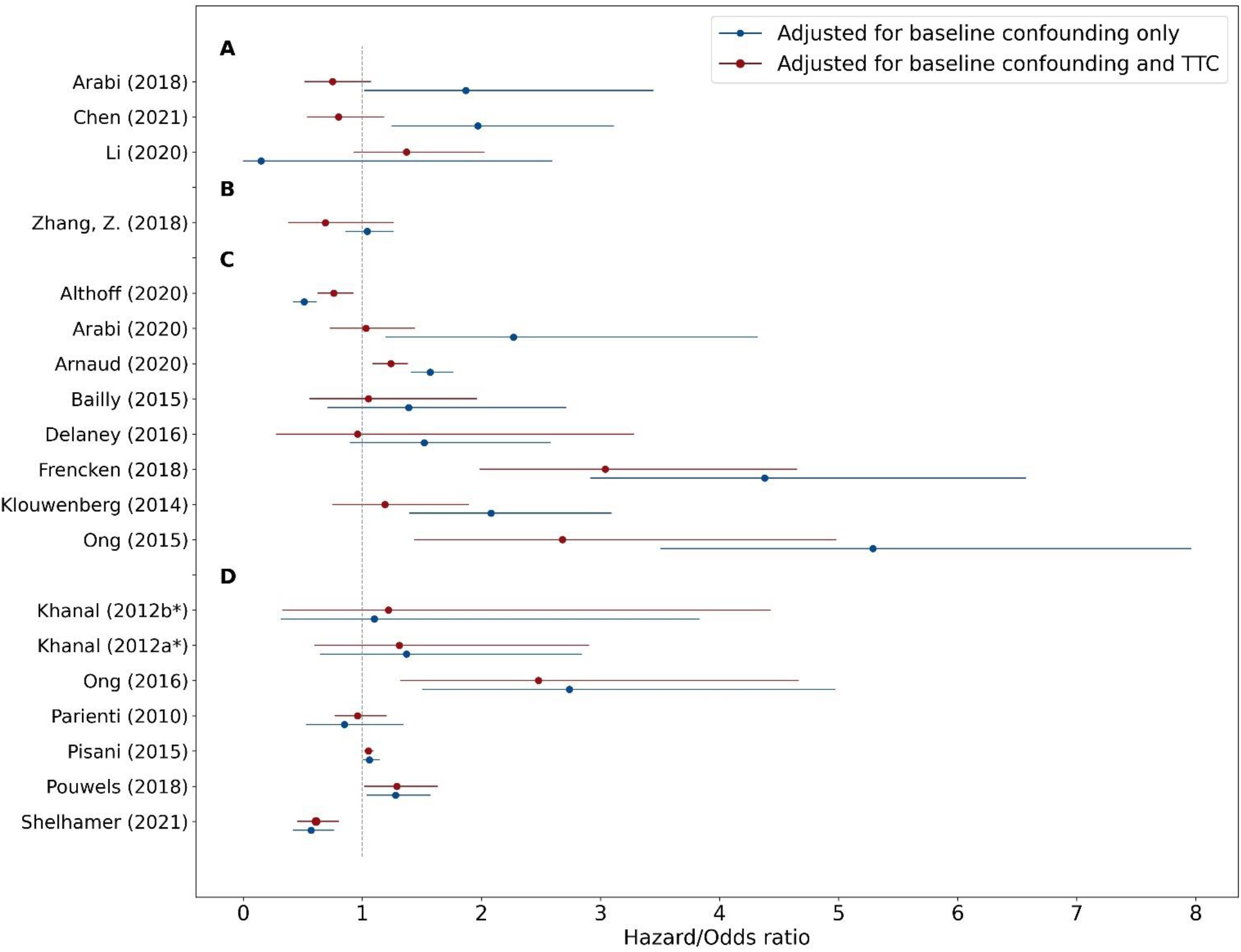
Difference in effect estimate in odds (ORs) or hazard ratios (HRs) by adjusting only for baseline confounding versus adjusting for baseline and treatment-affected time-varying confounding (TTC). In three studies (A), the point estimates moved to the opposite direction, in one (B) and seven (C) studies, the estimates moved away from and towards the null hypothesis, respectively. In six studies (D), there was a marginal difference in point estimates. Pouwels et al.^42^ estimated treatment effect on length-of-stay (expressed in days) by adjusting for baseline confounding and by adjusting for baseline confounding and TTC, and found a marginal difference in point estimates. *Khanal et al.^82^ compared prolonged intermittent renal replacement therapy with two different alternative treatments.

## Discussion

Our review of 60 published studies found a wide variety of treatments being studied, with a predominant focus on STRs, despite DTRs being most relevant in the ICU setting. We found incomplete reporting of the target trial components in most studies, among which the ‘treatment strategies’ and ‘analysis plan’ were incompletely reported most often. The causal assumptions often remained unmentioned, and this was especially noticeable in studies that applied RL methods.

ROBINS-I^53^ is a tool developed for assessing the risk of bias (ROB) in CI studies using observational data. Instead of assessing the ROB using this tool, we chose to assess the QOR. First, to fairly assess the ROB, the emulated target trial needs to be well reported, which was often not the case in the included studies (figure 3a). Moreover, ROB assessment would require expert knowledge of each specific treatment-outcome relationships studied in the included articles, which is beyond the scope of this review.

G methods and RL methods are often perceived as separate disciplines, but show great similarities. For example, Q-learning^54^ (an RL method, used by many of the included studies^46,47,55–57^) is very similar -and under certain conditions even algebraically equivalent- to G estimation (a G method).^58^ An important difference is that G methods are used for modelling both STRs and DTRs, while RL methods typically deal with DTRs. As both G methods and RL methods perform the same CI task (ie, finding optimal treatment regimes), both rely on the same, strong causal assumptions which should be acknowledged. While the consistency assumption is often plausible for treatments in the ICU, violations of the conditional exchangeability and positivity assumption are more likely and should be examined. Prior to examining violations of the causal assumptions, one needs a research question that is truly of interest in the ICU, a clear description of the target trial, and usage of a CI method that is appropriate for the type of studied treatment. The results of our review have led to five recommendations to improve future CI research and move towards actionable AI in the ICU (panel 2).

### Recommendations for future research

#### Ask the right research question

Treatments of interest in the ICU are typically DTRs and, therefore, this type of treatments is expected to be the focus of CI research in the ICU. However, 93% of the studies that used G methods studied STRs. To illustrate that many of these studies are considering research questions that are not truly of interest in the ICU, we will explore some examples. Zhang and colleagues[57] divided patients into two groups according to whether they received diuretics within the first two days of ICU admission or not. Thus, the emulated target trial answers the question whether or not to administer diuretics at the start of ICU admission. However, we argue that the question an ICU physician is really interested in is when to administer diuretics throughout the whole ICU stay, taking into account changing patient characteristics such as fluid balance (especially at later ICU stages). In addition, many of the included studies emulated target trials comparing ‘giving treatment sometime during follow-up’ versus ‘never giving treatment’. For example, Bailly and colleagues[52] studied the effect of systemic antifungal therapy, comparing a treated group (those who received antifungals during their ICU stay) with an untreated group (those who never received antifungals). As giving treatment ‘sometime during follow-up’ can be done in many ways, the estimated treatment effect is ill-defined and typically not truly of interest. In other words, both studies by Zhang[57] and Bailly[52] serve as examples of emulated RCTs that would never be conducted in the ICU.

#### Describe the question as a target trial emulation

To identify flaws in the relevance of a research question and correctness of the analysis, it is useful to describe the research question as a target trial emulation using the target trial framework^9^. Many of the included studies lacked a clear description of the ‘treatment strategies’ component of the target trial, that is, which treatment regimes are compared in the target trial. For example, Arabi and colleagues^59^ used IPTW to study the effect of corticosteroid therapy for ICU patients with Middle East Respiratory Syndrome. However, it remains unclear which treatment regimes (eg, ‘treat daily with corticosteroids’) are being compared. Moreover, roughly half of the included studies lacked a complete description of the ‘analysis plan’ component and therefore, are not reproducible. We advocate detailed description which allows reproduction of the used methodology, ideally accompanied with code and (example) data.

#### Use methods that suit the research question

We excluded 227 studies that modelled time-fixed treatments (figure 2). As time-fixed treatments in the ICU are rare, we hypothesize that in many of these studies, the implicit treatment of interest is time-varying. Research questions concerning time-varying treatments may be reformulated into simplified, time-fixed versions, because standard methods are easier to implement or high-quality, longitudinal data is unavailable. One may argue that, if the bias introduced by TTC^16,17^ is negligible, standard methods suffice for CI in time-varying treatments as well. However, empirical results from studies included in this review suggest that adjusting for TTC can lead to substantial differences in effect estimates and sometimes even to opposite conclusions (figure 4). Hence, it is possible that many of the excluded studies that implicitly studied time-varying treatments but modelled these as if they are time-fixed, published biased effect estimates. We advocate adjustment for TTC in any CI study where the treatment of interest is time-varying. Modelling DTRs is slightly more complex than STRs (which may be a reason for the focus on STRs among the included studies) and therefore requires different approaches. Various methods exist to find optimal DTRs, either from a set of pre-specified regimes or directly from data (for an overview, we refer to the book by Chakraborty and Moodie^60^). Among the included studies in this review, for example, Shahn and colleagues^31^ used ‘artificial censoring/IPW’^60–62^ to estimate the optimal fluid-limiting treatment regime for sepsis patients among a pre-specified set of DTRs (ie, ‘fluid caps’). Wang and colleagues^33^ used the parametric G formula to estimate the per-protocol (PP) effect of ‘low tidal volume ventilation’, a pre-specified DTR that was compared with standard care in an earlier RCT.^63^ Here, the target trial corresponds to the original RCT, but with full compliance. RL methods and G estimation can be used to approximate optimal DTRs without a pre-specified set of regimes. In RL studies, finding the optimal treatment regime (often referred to as the optimal ‘policy’, table S1) is typically followed by a validation step to quantify the value of the optimized regime (ie, OPE, panel 1). The ‘U-curve method’^38^ (a specific OPE method) is common among the included RL studies (figure S2) and is based on associating the difference between the (observed) clinician’s treatment regime and the (estimated) optimal treatment regime with patient outcome. As it completely ignores the potential effect of confounders, we recommend avoiding this method. We argue that G methods are essentially OPE methods and therefore, these could (and maybe should) be used to evaluate optimal treatment regimes found in RL studies.

#### Mind the conditional exchangeability assumption

Conditional exchangeability is never guaranteed using observational data as the absence of unmeasured confounders is not verifiable in the data. To think about residual confounding or selection bias, incorporation of subject-matter expertise is key. Causal diagrams (represented by directed acyclic graphs)^64,65^ are a visual way to represent this expert knowledge and can be useful to describe potential sources of bias. There are different approaches to quantify how potential violations of the conditional exchangeability would affect the study results.^66^ Indirect approaches consider, for instance, the effect of adding additional confounders.^9^ A ‘bias analysis’ (or sensitivity analysis)^28^ examines the characteristics of potential unmeasured confounders and can be useful to quantify how much bias it would produce as a function of those characteristics.

#### Mind the positivity assumption

The positivity assumption -on the contrary-is verifiable, although this is rather complex for time-varying treatments^29^ and, given its dynamic nature, violations are expected in the ICU setting. The intuition for this assumption is that one can only study a treatment regime using data of patients who have received treatment that conform to this regime. The number of patient treatment histories that match the treatment regime of interest (ie, the ‘effective sample size’^67^) shrinks with the number of treatment decisions in the patient’s history (which tends to be high in the ICU). For example, Gottesman and colleagues^38^ applied RL to a dataset of 3,855 patients to find an optimal treatment regime for sepsis, but the effective sample size for this regime was only a few dozen. A small effective sample size makes positivity violations likely and leads to high uncertainties in estimated treatment effects. A straight-forward opportunity to tackle this challenge is increasing the sample size. Therefore, we advocate more usage (if appropriate) of the four currently available open-source ICU databases.^68^ However, increasing the sample size does not guarantee increasing the *effective* sample size, as the patients in the extra dataset may not be treated according to the regime of interest. Hence, another opportunity to increase the effective sample size is to minimize the mismatches between the treatment regime(s) of interest and those observed in the data, for instance, by avoiding modelling treatment regimes which differ greatly from the treatment protocol in place.

To detect (but not rule out) violations of the positivity assumption, examination of the distribution of the estimated (stabilized) IPT weights can be useful.^29^ This was common among the included studies that used IPTW (n=16/36, table S7), but is recommended in studies that use other CI methods as well. For RL studies that use an importance-sampling^69^ OPE method, it is recommended to examine the distribution of importance weights^38^ (which is closely related to examination of IPT weights).

In studies using IPTW, weight stabilization and truncation can be used to limit high uncertainties in the effect estimates. Weight stabilization can improve the precision of effect estimates without the introduction of bias. However, a model based on stabilized weights results in a slightly different effect estimate compared to non-stabilized weights^70^ and should be interpreted carefully. Weight truncation also improves precision, but at the expense of bias. Examination of the influence of the introduced bias by checking the change of the effect estimates under progressive truncation of IP weights is recommended.^25^

### Study limitations

First, whereas efforts were made to ensure that the literature search was comprehensive, we could have missed studies for different reasons. Some research may have used non-conventional terminology to describe the used CI method, or used a CI method which was not included in our search strategy. For example, dynamic weighted ordinary least squares (dWOLS)^71,72^ is a relatively new method which has been used to model DTRs in the ICU setting in several studies.^73,74^ This method benefits from properties of both Q-learning (an RL method) and G-estimation (a G method) and may be an interesting direction for future research. Second, only one reviewer (JS) performed title-abstract screening and item collection, although thorough discussions with the other reviewers (MvG, JK, JL) occurred in case of uncertainty. Third, items that were not collected could be of interest for future investigation. For example, we did not differentiate RL further into specific RL methods.

### Study strengths

This systematic review stresses the importance of causality for actionable AI and provides a contemporary overview of CI research in the ICU literature. We describe shortcomings of the identified studies in terms of reporting and, furthermore, provide handles for improving future CI research. These recommendations are not limited to the ICU but apply to medical CI research as a whole. Unlike other systematic reviews on time-varying medical treatments,^75–77^ we did not limit our focus to either G methods or RL, but rather acknowledge that both these method classes can be used to perform CI tasks and therefore, hold the promise to bring actionable AI to the bedside.

### Conclusion

Towards actionable AI in the ICU, we concur with the guidance of editors of critical care journals^78,79^ to change the focus of observational research in the ICU from prediction to CI. To unlock this potential in a trustworthy and responsible manner, we advocate development of models for CI focusing on clinically relevant treatments, using a description of the research question as a target trial emulation, choosing appropriate CI methods given the treatment of interest and acknowledging (and ideally examining potential violations) of the causal assumptions.

### Panel 2: Summary of recommendations for future research

#### Ask the right research question

When developing a model for CI, consider clinically relevant treatments. In the ICU, treatment decisions typically occur at multiple time point during admission (ie, time-varying treatments) and often depend on the patient’s response to treatment (ie, dynamic treatment regimes).

#### Describe the question as a target trial emulation

To identify flaws in the relevance of a research question and correctness of the analysis, it is useful to imagine a randomized trial that would have answered the research question (ie, the target trial), describe its components using the target trial framework and emulate it to the extent possible.

#### Use methods that suit the research question

Standard methods (like regression) are easy to implement and suffice for time-fixed treatments, but lead to biased estimates when used for time-varying treatments. Therefore, adjustment for bias introduced by treatment-affected time-varying confounding is always recommended when dealing with time-varying treatments. Modelling of dynamic treatment regimes requires slightly different approaches compared to static treatment regimes.

#### Mind the conditional exchangeability assumption

CI is not possible based on data only, and incorporation of expert knowledge is key to think about the causal structure between the treatment and outcome of interest. Representing this expert knowledge in causal diagrams is useful to visualize potential sources of bias. Although violations of this assumption can never be completely ruled out using observational data, several approaches exist to examine potential violations. For example, a bias analysis can be helpful to quantify how much bias unmeasured confounders could produce.

#### Mind the positivity assumption

This assumption is verifiable, but this is rather complex for time-varying treatments and violations are expected given the dynamic nature of the ICU. Violations could be minimized by increasing the sample size (eg, by more usage of open-source ICU databases) and studying treatment regimes that are similar to those observed in the data. Examination of estimated inverse-probability-of-treatment weights is useful to detect (but not rule out) positivity violations.

## Data Availability

All data produced in the present work are contained in the manuscript.

### Appendix A: Embase search strategy

#### Embase.com

(‘causal inference’/de OR ‘causal model’/de OR ‘causal modeling’/de OR ‘inverse probability weighting’/de OR ((causal NEAR/3 (inferen* OR model*)) OR ((causal OR average-treatment* OR individuali*-treatment* OR personali*-treatment*) NEXT/1 (effect*)) OR time-vary*-confound* OR g-computation* OR g-estimation* OR g-formula* OR doubly-robust OR counterfactual* OR (inverse-probabilit* NEAR/3 (weight* OR estimat*)) OR ((marginal-structur* OR structural-nest* OR causal-effect* OR causal-graphic* OR causal-inferen* OR condition*-outcome* OR sequen*-cox*) NEAR/3 (method* OR model*)) OR TAR-Net OR (Treatment*-Agnost* NEAR/3 Representat* NEAR/3 Network*) OR double-machine-learning OR anchor*-regress* OR x-learner* OR t-learner* OR s-learner* OR q-learning OR q-network OR reinforcement*-learn* OR ((policy OR value) NEXT/1 iteration*) OR temporal-differen* OR actor-critic* OR (Markov NEAR/3 decision NEAR/3 process*)):ab,ti) AND (‘intensive care’/exp OR ‘intensive care unit’/exp OR ‘critically ill patient’/de OR ‘critical illness’/de OR ‘artificial ventilation’/exp OR ‘mechanical ventilator’/exp OR (intensive-care* OR critical-care* OR critical*-ill* OR icu OR ((mechanic* OR artificial*) NEAR/3 ventilat*)):Ab,ti,jt) NOT [conference abstract]/lim AND [english]/lim NOT (‘pediatric intensive care unit’/de OR ‘neonatal intensive care unit’/de OR child/exp OR pediatrics/exp OR (nicu OR picu OR nicus OR picus OR infant* OR child* OR neonat* OR newborn* OR pediatr* OR paediatr*):ab,ti)

#### Medline ALL

(((caus* ADJ3 (inferen* OR model*)) OR ((causal OR average-treatment* OR individuali*-treatment* OR personali*-treatment*) ADJ (effect* OR method*)) OR time-vary*-confound* OR g-computation* OR g-estimation* OR g-formula* OR doubly-robust-estimation* OR counterfactual* OR (inverse-probabilit* ADJ3 (weight* OR estimat*)) OR ((marginal-structur* OR structural-nest* OR causal-effect* OR causal-graphic* OR causal-inferen* OR semi-paramet* OR semiparamet* OR fully-paramet*) ADJ3 (method* OR model*)) OR TAR-Net OR (Treatment*-Agnost* ADJ3 Representat* ADJ3 Network*) OR double-machine-learning OR anchor*-regress* OR x-learner* OR t-learner* OR s-learner* OR q-learning OR q-network OR reinforcement*-learn* OR ((policy OR value) ADJ iteration*) OR temporal-differen* OR actor-critic* OR (Markov ADJ3 decision ADJ3 process*)).ab,ti. OR (RL OR IRL).ti.) AND (exp Intensive Care Units/ OR Critical Illness/ OR exp Respiration, Artificial/ OR exp Ventilators, Mechanical/ OR (intensive-care* OR critical-care* OR critical*-ill* OR icu OR ((mechanic* OR artificial*) ADJ3 ventilat*)).ab,ti,jt) NOT (conference abstract) AND english.la. NOT (Intensive Care Units, Pediatric/de OR Intensive Care Units, Neonatal/de OR exp Child/ OR exp pediatrics/ OR (nicu OR picu OR nicus OR picus OR infant* OR child* OR neonat* OR newborn* OR pediatr* OR paediatr*).ti,ab)

#### Web of Science Core Collection

TS=(((causal NEAR/2 (inferen* OR model*)) OR ((causal OR average-treatment* OR individuali*-treatment* OR personali*-treatment*) NEAR/1 (effect*)) OR time-vary*-confound* OR g-computation* OR g-estimation* OR g-formula* OR doubly-robust OR counterfactual* OR (inverse-probabilit* NEAR/2 (weight* OR estimat*)) OR ((marginal-structur* OR structural-nest* OR causal-effect* OR causal-graphic* OR causal-inferen* OR condition*-outcome* OR sequen*-cox*) NEAR/2 (method* OR model*)) OR TAR-Net OR (Treatment*-Agnost* NEAR/2 Representat* NEAR/2 Network*) OR double-machine-learning OR anchor*-regress* OR x-learner* OR t-learner* OR s-learner* OR q-learning OR q-network OR reinforcement*-learn* OR ((policy OR value) NEAR/1 iteration*) OR temporal-differen* OR actor-critic* OR (Markov NEAR/2 decision NEAR/2 process*)) AND (intensive-care* OR critical-care* OR critical*-ill* OR icu OR ((mechanic* OR artificial*) NEAR/2 ventilat*)) NOT (nicu OR picu OR nicus OR picus OR infant* OR child* OR neonat* OR newborn* OR pediatr* OR paediatr*)) AND DT=(Article OR Review OR Letter OR Early Access)

#### Google Scholar

Searched with 2 different queries:

- “causal inference”|”marginal structural models”|”g-formula”|”structural nested models”|”reinforcement learning” “intensive|critical care” Only the <u>first 200</u> results
- “causal inference”|”marginal structural models”|”g-formula”|”structural nested models”|”reinforcement learning” intitle:”intensive|critical care” <u>All 389</u> results

#### MedRxiv and BioRxiv

searched via Google with the following query:

inurl:medrxiv|biorxiv filetype:pdf “causal inference”|”marginal structural models”|”g-formula”|”structural nested models”|”reinforcement learning” “intensive|critical care”

When: 2 March 2022

Settings:

SafeSearch Filters turned on

Auto-complete with trending searches: Show popular searches

Region Settings: Current region (i.e. the Netherlands)

### Appendix C: Supplementary figures

**Figure S1:**
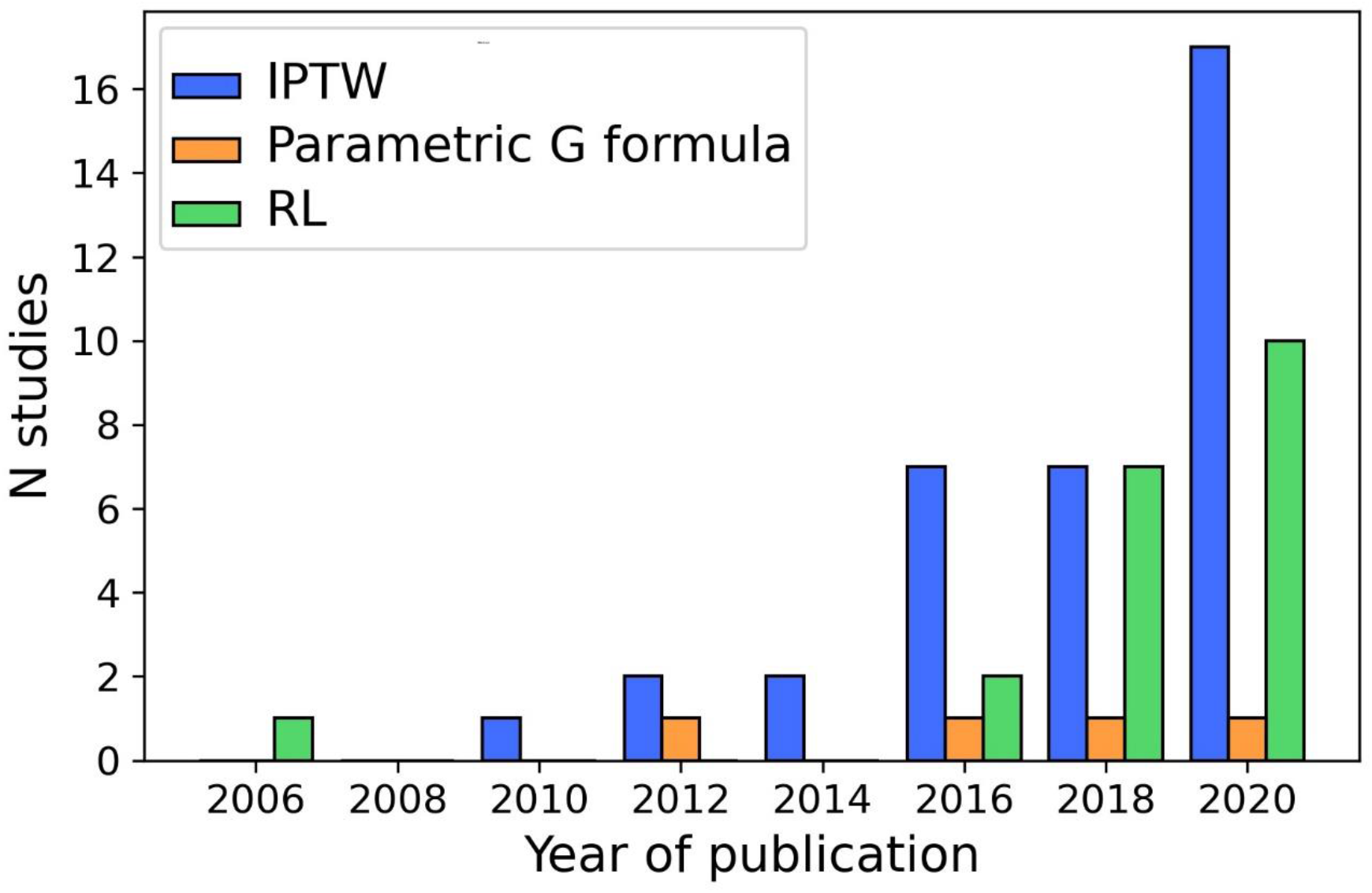
Bar chart representing the number of published articles using the different modelling strategies over the years.

**Figure S2:**
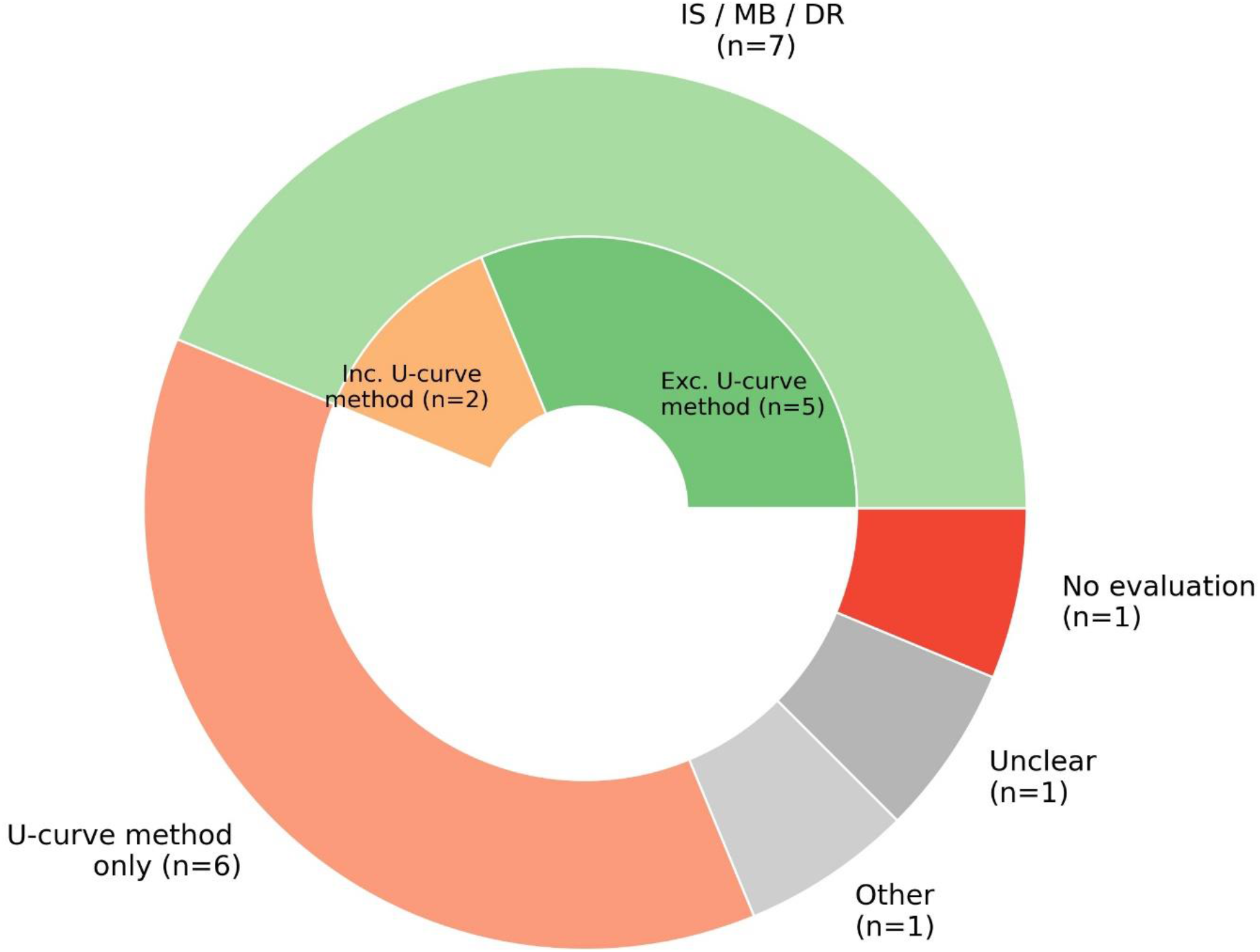
Nested pie chart representing the off-policy evaluation (OPE) methods used in the reinforcement learning studies that used real patient data (n=16). IS=Importance sampling, MB=Model-based, DR=Doubly robust.

**Figure S3:**
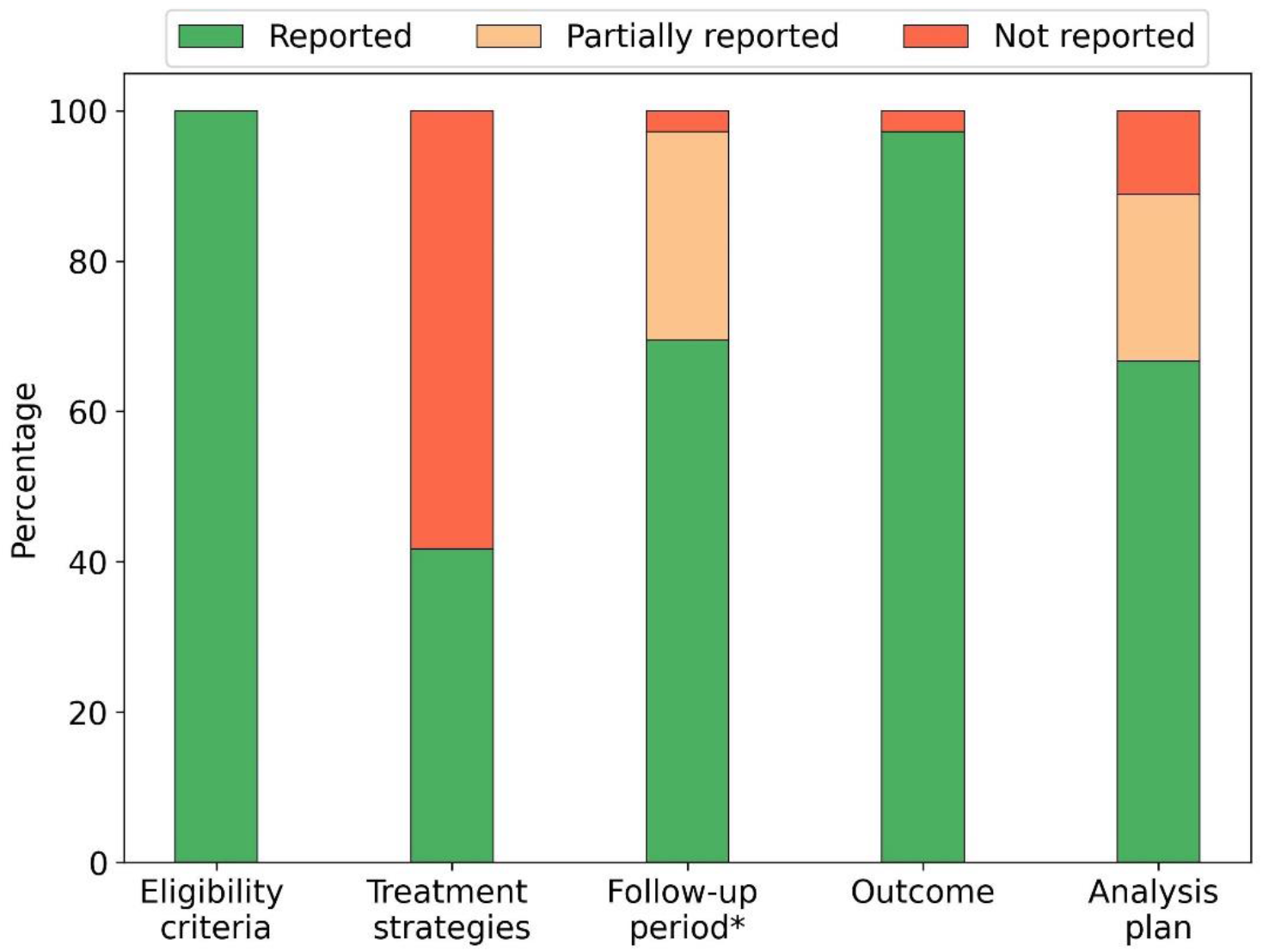
Reporting of the target trial components in studies using IPTW (n=36).

**Figure S4:**
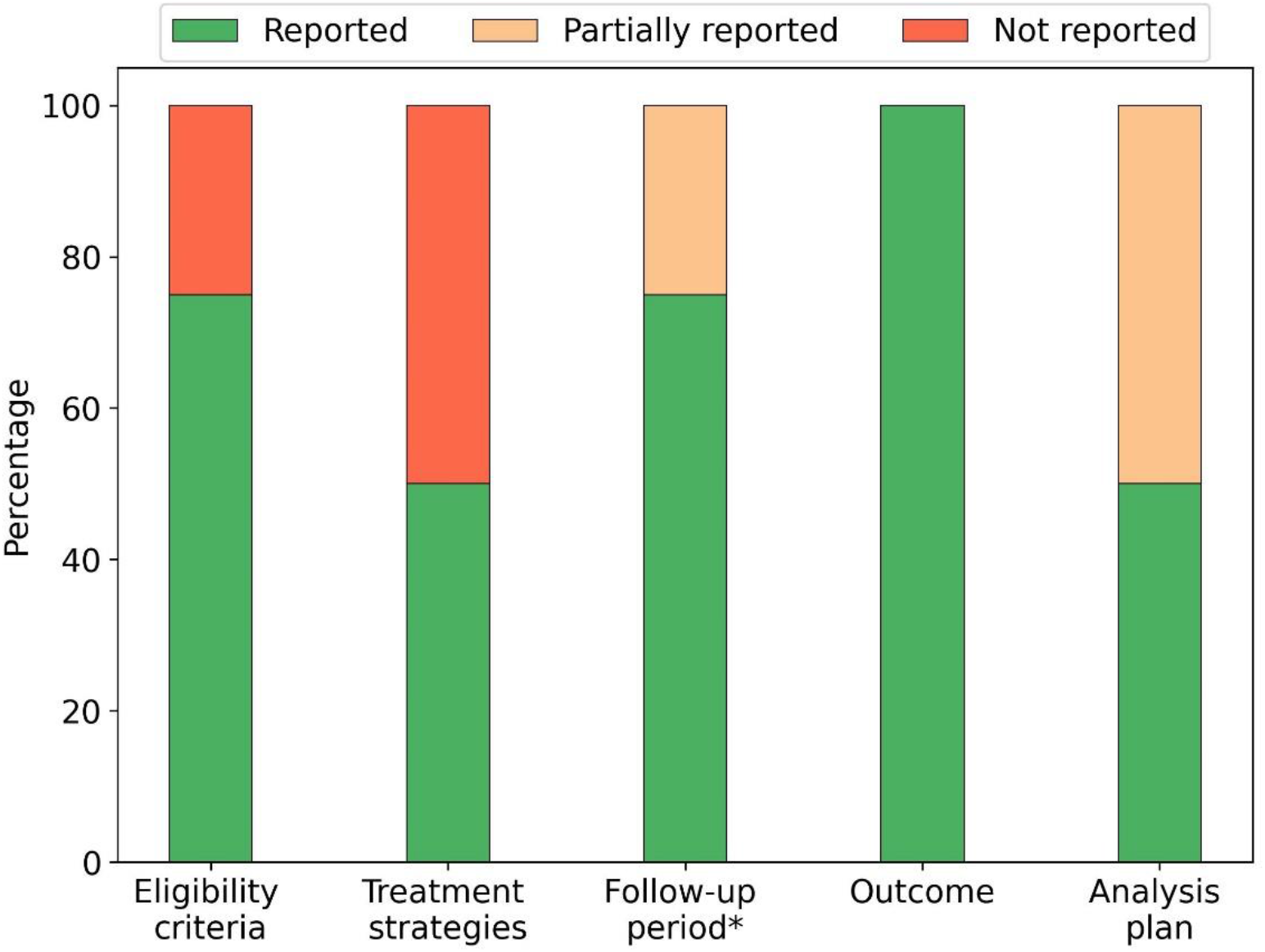
Reporting of the target trial components in studies using the parametric G formula (n=4).

**Figure S5:**
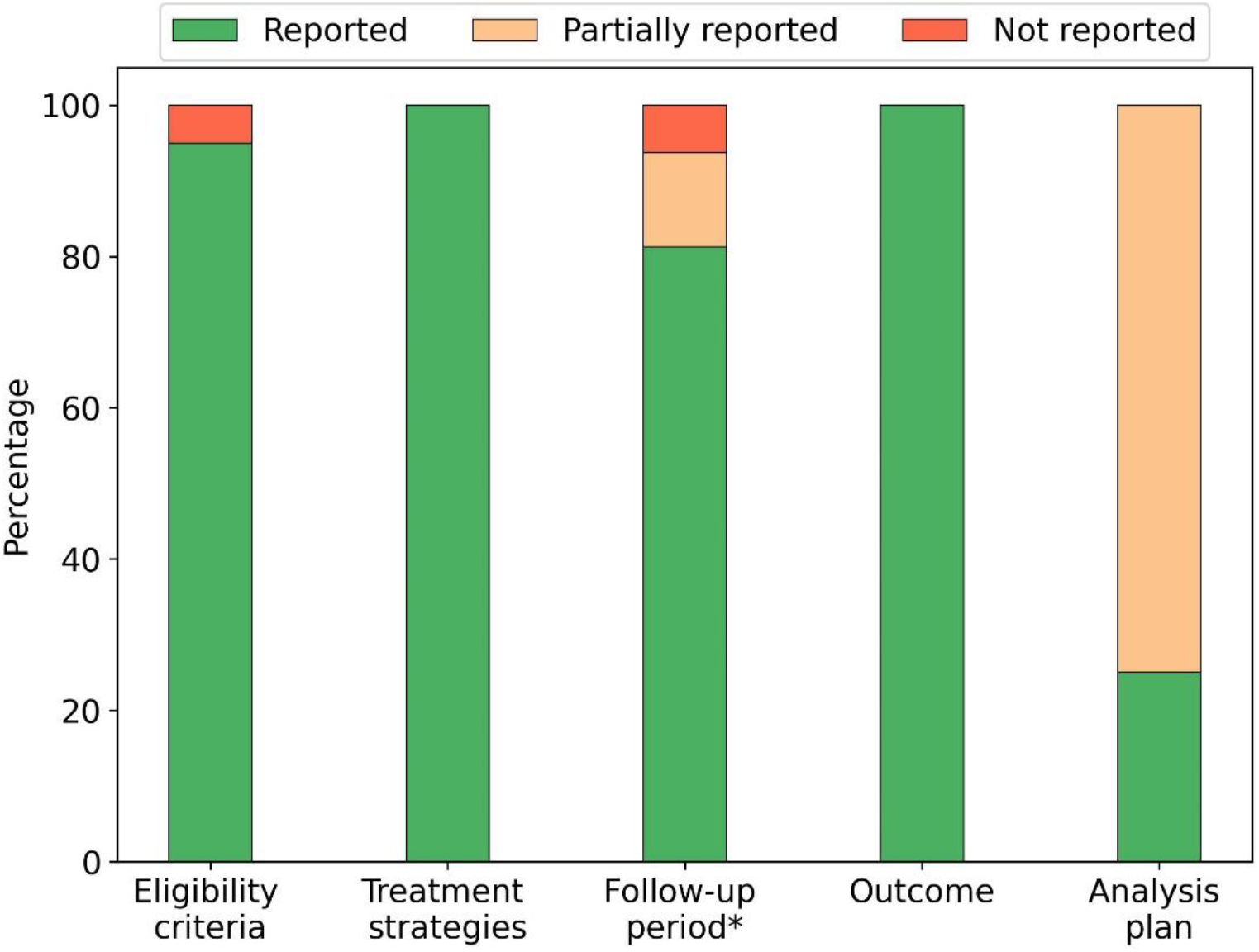
Reporting of the target trial components in studies using RL (n=20). *For the follow-up component, the studies that used simulated patient data (n=4) are not taken into account.

**Figure S6:**
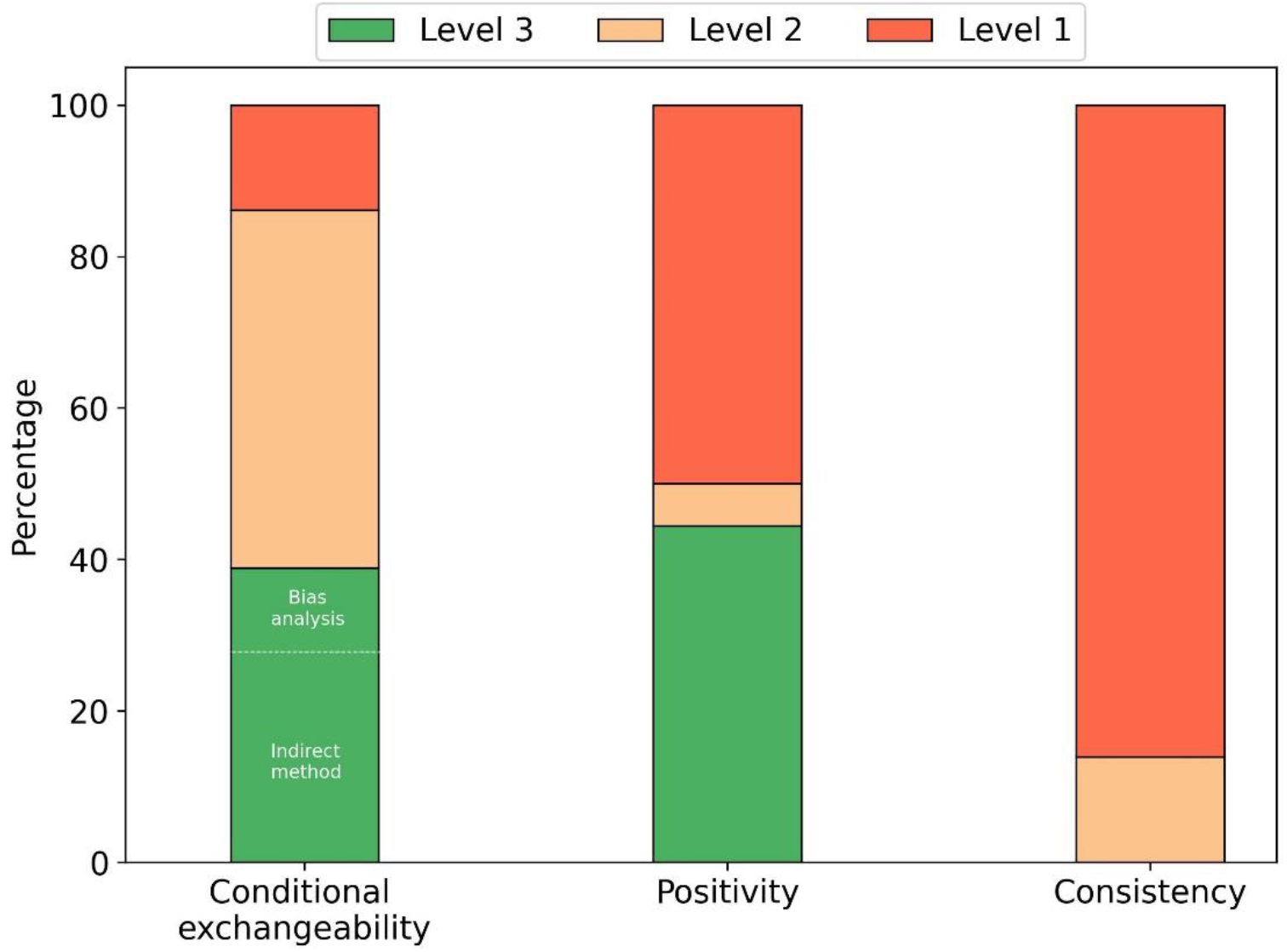
Reporting of assumptions in the studies using IPTW (n=36). Level 1=assumption not mentioned, level 2=assumption mentioned, level 3=attempt to check for potential violations of the assumption reported.

**Figure S7:**
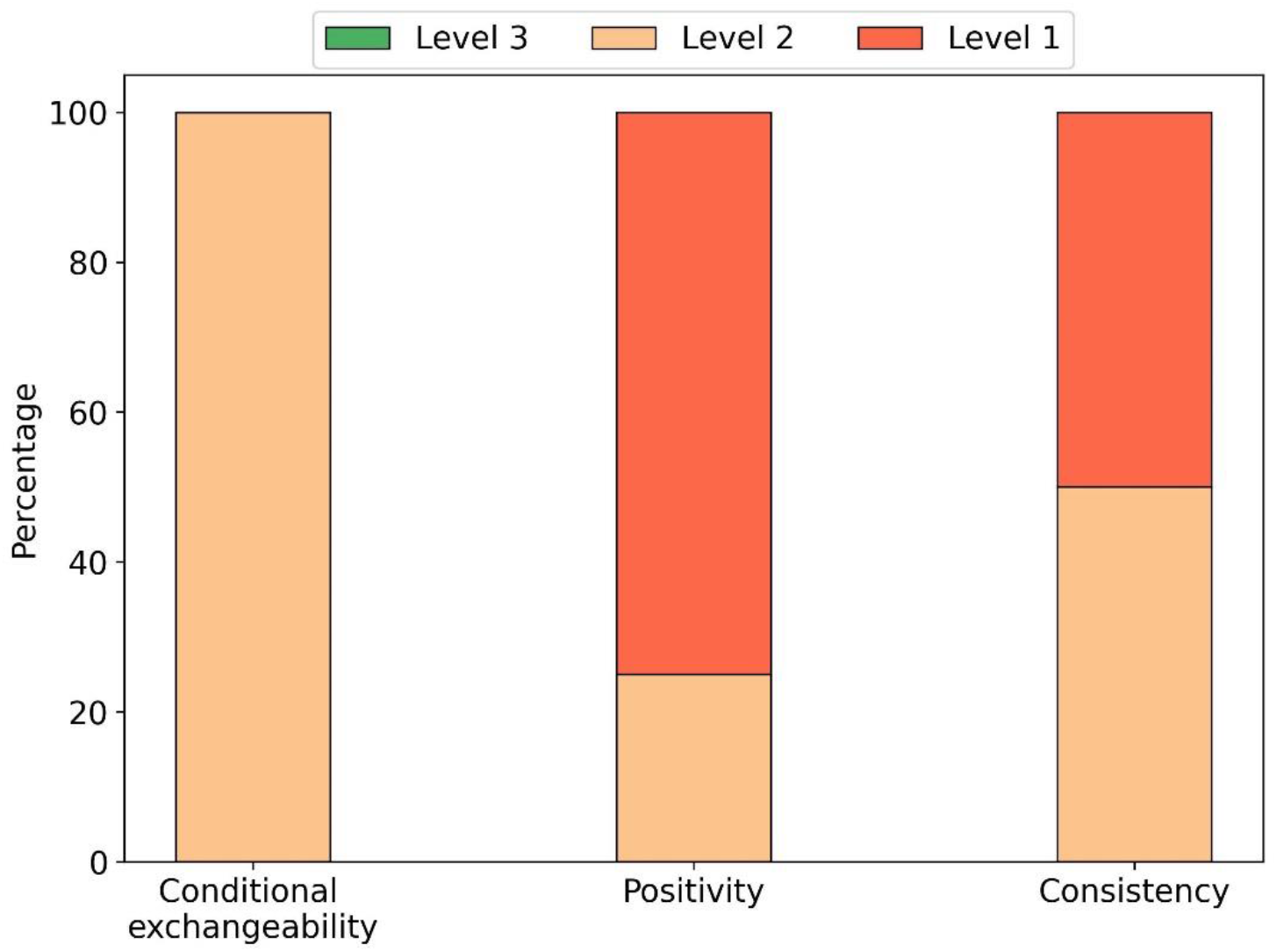
Reporting of assumptions in the studies using the parametric G formula (n=4). Level 1=assumption not mentioned, level 2=assumption mentioned, level 3=attempt to check for potential violations of the assumption reported.

**Figure S8:**
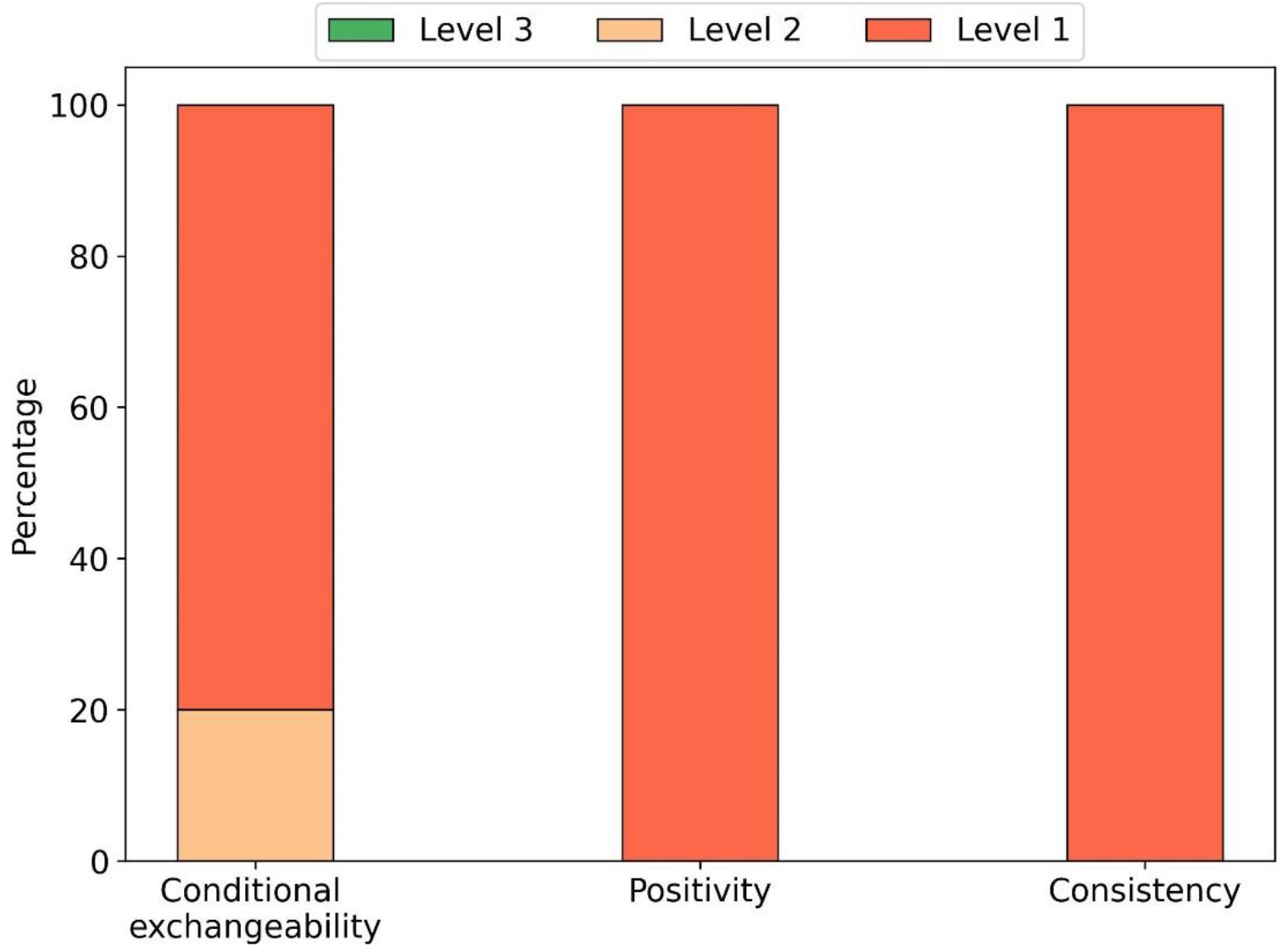
Reporting of assumptions in the studies using RL (n=20). Level 1=assumption not mentioned, level 2=assumption mentioned, level 3=attempt to check for potential violations of the assumption reported.

### Appendix D: Supplementary tables

**Table S1:** Commonly used terms (not synonyms) to describe similar concepts typically used in research using G methods and RL methods.

| **G methods** | **Reinforcement learning** | **Other commonly used terms** |
| --- | --- | --- |
| Treatment | Action | Exposure, intervention |
| Outcome | Reward |  |
| (Treatment) regime | Policy | strategy, regimen, decision rule, joint exposures, sustained strategy, plan, protocol |
| Structural causal model | Environment model | World model |
| (Conditional) exchangeability | Unconfoundedness | Ignorability, no unmeasured/residual confounding |
| Positivity | Feasibility | Experimental treatment assignment, common support, overlap |
| Observational data | Data collected under clinician's policy |  |

**Table S2:** Subcomponents of the analysis plan component specifically for each used CI method.

| **IPTW** | **Parametric G formula** | **RL** |
| --- | --- | --- |
| Propensity score estimator | Outcome estimator | Learning scheme |
| Propensity score predictors | Outcome predictors | State space model |
|  | Confounders estimators | Environment model |
|  | Confounders predictors | Discount factor |
|  | Method to evaluate the G formula |  |

**Table S3:** List with collected items per study. NA=not applicable

| *Reference* | **Treatment of interest (categorized)** | **Primary outcome (categorized)** | **Number of included ICUs** | **Usage of open source databases** | **Study size (n patients)** | **Treatment strategy type** | **Modelling strategy** |
| --- | --- | --- | --- | --- | --- | --- | --- |
| *Agodi 2017* | Protocol compliance | Nosocomial infections | 17 | None | 1840 | Static | parametric G formula |
| *Althoff 2020* | NIV | Need for MV | 682 | None | 53654 | Static | IPTW |
| *Amer 2021* | Anti-inflammatory drugs | VFDs | 168 | None | 860 | Static | IPTW |
| *Arabi 2018* | Anti-inflammatory drugs | Mortality | 14 | None | 309 | Static | IPTW |
| *Arabi 2020* | Antimicrobials | Mortality | 14 | None | 349 | Static | IPTW |
| *Arnaud 2020* | Antimicrobials | AKI | 1 | MIMIC-III | 26865 | Static | IPTW |
| *Bailly 2015* | Antimicrobials | Mortality | 5 | None | 1491 | Static | IPTW |
| *Bekaert 2011* | Nosocomial infections | Mortality | 17 | None | 4479 | Static | IPTW |
| *Chen 2021* | Anti-inflammatory drugs | Mortality | 1 | None | 428 | Static | IPTW |
| *Cheng 2019* | Ordering of labs | Combined | 1 | MIMIC-III | 6060 | Dynamic | RL |
| *De Bus 2020* | Antimicrobials | Clinical cure | 152 | None | 1495 | Static | IPTW |
| *Delaney 2016* | Anti-inflammatory drugs | Mortality | 51 | None | 607 | Static | IPTW |
| *Dupuis 2017* | Blood transfusion | Mortality | 23 | None | 6016 | Static | IPTW |
| *Eghbali 2021* | Sedatives & analgesics | Maintenance of clinical target value | 1 | MIMIC-IV | 1757 | Dynamic | RL |
| *Esperatti 2013* | Multiple | Mortality | 6 | None | 335 | Static | IPTW |
| *Frencken 2018* | Bacterial colonization | Nosocomial infections | 1 | None | 2066 | Static | IPTW |
| *Jeter 2021* | Vasopressors & IV fluids | Combined | 1 | MIMIC-III | 5366 | Dynamic | RL |
| *Khanal 2012* | RRT | Mortality | 1 | None | 146 | Static | IPTW |
| *Klouwenberg 2014* | Nosocomial infections | Mortality | 1 | None | 1112 | Static | IPTW |
| *Komorowski 2018* | Vasopressors & IV fluids | Mortality | 209 | MIMIC-III + eICU | 96156 | Dynamic | RL |
| *Lal 2020* | Vasopressors & IV fluids | Other | 1 | None | 29 | Dynamic | Digital Twin |
| *Lejarza 2021* | Discharge from ICU | Successful discharge | 1 | MIMIC-III | 19664 | Dynamic | RL |
| *Li 2019* | gastric acid-suppressing agents | Nosocomial infections | 1 | None | 6133 | Static | parametric G formula |
| *Li 2020* | Anti-inflammatory drugs | Mortality | 10 | None | 294 | Static | IPTW |
| *Libório 2020* | Diuretics | Mortality | 1 | MIMIC-III | 14896 | Static | IPTW |
| *Lin 2018* | Anticoagulants | Maintenance of clinical target value | 1 | MIMIC-III | 4908 | Dynamic | RL |
| *Liu 2016* | Multiple | Vital signs | 1 | None | 300 | Static | parametric G formula |
| *Lopez-Martinez 2019* | Sedatives & analgesics | Combined | 1 | MIMIC-III | 6843 | Dynamic | RL |
| *Mecklenburg 2021* | therapeutic hypothermia (TH) | Major bleeding | 1 | None | 66 | Static | IPTW |
| *Muriel 2015* | Sedatives & analgesics | Need for MV | 322 | None | 842 | Static | IPTW |
| *Nemati 2016* | Anticoagulants | Maintenance of clinical target value | 1 | MIMIC-II | 4470 | Dynamic | RL |
| *Ohbe 2018* | Nutrition | Mortality | 1200 | None | 1769 | Static | IPTW |
| *Ong 2015* | Nosocomial infections | Mortality | 2 | None | 3080 | Static | IPTW |
| *Ong 2016* | Nosocomial infections | Mortality | 2 | None | 399 | Static | IPTW |
| *Padmanabhan 2015* | Sedatives & analgesics | Deviation from vital signs target value | NA | None | NA | Dynamic | RL |
| *Padmanabhan 2017* | Sedatives & analgesics | Maintenance of clinical target value | NA | None | NA | Dynamic | RL |
| *Parienti 2010* | RRT | Bacterial colonization | 12 | None | 736 | Static | IPTW |
| *Peine 2021* | MV | Mortality | 209 | MIMIC-III + eICU | 37029 | Dynamic | RL |
| *Petersen 2019* | Anti-inflammatory drugs | Mortality | NA | None | NA | Dynamic | RL |
| *Pisani 2015* | antipsychotic | Delirium | 1 | None | 93 | Static | IPTW |
| *Pouwels 2017* | Nosocomial infections | Mortality | 2 | None | 3411 | Static | IPTW |
| *Pouwels 2018* | Nosocomial infections | Mortality | 2 | None | 2914 | Static | IPTW |
| *Pouwels 2020* | Nosocomial infections | ICU LOS | 2 | None | 2914 | Static | IPTW |
| *Prasad 2017* | MV | Combined | 1 | MIMIC-III | 8182 | Dynamic | RL |
| *Raghu 2017* | Vasopressors & IV fluids | Combined | 1 | MIMIC-III | 17898 | Dynamic | RL |
| *Raghu 2018* | Vasopressors & IV fluids | Combined | 1 | MIMIC-III | 17898 | Dynamic | RL |
| *Roggeveen 2021* | Vasopressors & IV fluids | Mortality | 2 | MIMIC-III + AmsterdamUMCdb | 11382 | Dynamic | RL |
| *Shahn 2020* | Vasopressors & IV fluids | Mortality | 1 | MIMIC-III | 1639 | Dynamic | IPTW |
| *Shahn 2021* | Vasopressors & IV fluids | Mortality | 1 | MIMIC-III | 1639 | Dynamic | IPTW |
| *Shelhamer 2021* | Prone positioning | Mortality | 1 | None | 335 | Static | IPTW |
| *Sinzinger 2005* | Sedatives & analgesics | Maintenance of clinical target value | NA | None | NA | Dynamic | RL |
| *Steen 2021* | Nosocomial infections | Mortality | 1 | None | 2720 | Static | IPTW |
| *Sun 2021* | vital signs | Combined | 1 | MIMIC-III | 28840 | Dynamic | RL |
| *Tacquard 2021* | Anticoagulants | Thrombotic complications | 8 | None | 538 | Static | IPTW |
| *Truche 2016* | RRT | Mortality | 19 | None | 1360 | Static | IPTW |
| *Wang 2011* | MV | Mortality | 23 | None | 1410 | Dynamic | parametric G formula |
| *Weng 2017* | vital signs | Mortality | 1 | MIMIC-III | 5565 | Dynamic | RL |
| *Zhang, R. 2021* | Diuretics | Mortality | 20 | None | 932 | Static | IPTW |
| *Zhang, Z. 2018* | Vasopressors & IV fluids | Mortality | 1 | MIMIC-III | 1718 | Static | IPTW |
| *Zhang, Z. 2019* | Vasopressors & IV fluids | Mortality | 1 | MIMIC-III | 3406 | Static | IPTW |
| *Zheng 2021* | NIV | Mortality | 1 | None | 1372 | Dynamic | RL |

**Table S4:** Subcomponent-specific results of the quality of reporting assessment in the reproducibility domain, specifically for the studies using IPTW.

| Reference | Eligibility criteria | Treatment strategies | Outcome | Follow-up |  |  | Analysis plan |  |
| --- | --- | --- | --- | --- | --- | --- | --- | --- |
|  |  |  |  | Time zero | End of follow-up | Time resolution | Propensity score estimator | Propensity score predictors |
| Althoff 2020 | 😊 | 😞 | 😊 | 😊 | 😊 | 😊 | 😞 | 😊 |
| Amer 2021 | 😊 | 😞 | 😊 | 😊 | 😊 | 😊 | 😞 | 😊 |
| Arabi 2018 | 😊 | 😞 | 😊 | 😊 | 😊 | 😊 | 😞 | 😊 |
| Arabi 2020 | 😊 | 😞 | 😊 | 😊 | 😊 | 😊 | 😞 | 😊 |
| Arnaud 2020 | 😊 | 😞 | 😊 | 😊 | 😊 | 😊 | 😊 | 😊 |
| Bailly 2015 | 😊 | 😞 | 😊 | 😊 | 😊 | 😊 | 😊 | 😊 |
| Bekaert 2011 | 😊 | 😊 | 😊 | 😊 | 😊 | 😊 | 😊 | 😊 |
| Chen 2021 | 😊 | 😞 | 😊 | 😊 | 😊 | 😊 | 😞 | 😊 |
| De Bus 2020 | 😊 | 😊 | 😊 | 😊 | 😊 | 😊 | 😊 | 😊 |
| Delaney 2016 | 😊 | 😞 | 😊 | 😊 | 😊 | 😊 | 😊 | 😊 |
| Dupuis 2017 | 😊 | 😞 | 😊 | 😊 | 😊 | 😊 | 😊 | 😊 |
| Esperatti 2013 | 😊 | 😞 | 😊 | 😊 | 😊 | 😞 | 😊 | 😊 |
| Frencken 2018 | 😊 | 😞 | 😊 | 😊 | 😊 | 😊 | 😞 | 😊 |
| Khanal 2012 | 😊 | 😞 | 😊 | 😞 | 😊 | 😊 | 😊 | 😊 |
| Klouwenberg 2014 | 😊 | 😊 | 😊 | 😞 | 😞 | 😊 | 😊 | 😊 |
| Li 2020 | 😊 | 😞 | 😊 | 😞 | 😞 | 😊 | 😞 | 😞 |
| Libório 2020 | 😊 | 😞 | 😊 | 😊 | 😊 | 😊 | 😊 | 😊 |
| Mecklenburg 2021 | 😊 | 😊 | 😊 | 😊 | 😊 | 😞 | 😊 | 😊 |
| Muriel 2015 | 😊 | 😞 | 😞 | 😊 | 😊 | 😞 | 😊 | 😊 |
| Ohbe 2018 | 😊 | 😊 | 😊 | 😞 | 😞 | 😊 | 😊 | 😊 |
| Ong 2015 | 😊 | 😊 | 😊 | 😊 | 😊 | 😊 | 😊 | 😞 |
| Ong 2016 | 😊 | 😊 | 😊 | 😊 | 😊 | 😊 | 😊 | 😊 |
| Parienti 2010 | 😊 | 😞 | 😊 | 😞 | 😞 | 😞 | 😞 | 😞 |
| Pisani 2015 | 😊 | 😞 | 😊 | 😊 | 😊 | 😊 | 😊 | 😊 |
| Pouwels 2017 | 😊 | 😞 | 😊 | 😞 | 😞 | 😊 | 😊 | 😊 |
| Pouwels 2018 | 😊 | 😊 | 😊 | 😞 | 😞 | 😊 | 😊 | 😊 |
| Pouwels 2020 | 😊 | 😊 | 😊 | 😊 | 😊 | 😊 | 😊 | 😊 |
| Shahn 2020 | 😊 | 😊 | 😊 | 😊 | 😊 | 😊 | 😊 | 😊 |
| Shahn 2021 | 😊 | 😊 | 😊 | 😊 | 😞 | 😊 | 😊 | 😊 |
| Shelhamer 2021 | 😊 | 😞 | 😊 | 😊 | 😞 | 😞 | 😞 | 😞 |
| Steen 2021 | 😊 | 😊 | 😊 | 😊 | 😊 | 😊 | 😊 | 😊 |
| Tacquard 2021 | 😊 | 😞 | 😊 | 😊 | 😊 | 😞 | 😞 | 😞 |
| Truche 2016 | 😊 | 😞 | 😊 | 😊 | 😞 | 😊 | 😊 | 😊 |
| Zhang, R. 2021 | 😊 | 😞 | 😊 | 😞 | 😊 | 😊 | 😞 | 😞 |
| Zhang, Z. 2018 | 😊 | 😞 | 😊 | 😞 | 😞 | 😊 | 😞 | 😊 |
| Zhang, Z. 2019 | 😊 | 😞 | 😊 | 😞 | 😞 | 😊 | 😊 | 😊 |

**Table S5:** Subcomponent-specific results of the quality of reporting assessment in the reproducibility domain, specifically for the studies using the parametric G formula.

| Reference | Eligibility criteria | Treatment strategies | Outcome | Follow up |  |  | Analysis plan |  |  |  |  |
| --- | --- | --- | --- | --- | --- | --- | --- | --- | --- | --- | --- |
|  |  |  |  | Time zero | End of follow-up | Time resolution | Outcome estimator | Outcome predictors | Confounders estimators | Confounders predictors | Method to evaluate the G formula |
| Agodi 2017 | 😊 | 😞 | 😊 | 😊 | 😊 | 😞 | 😊 | 😞 | 😞 | 😞 | 😊 |
| Li 2019 | 😊 | 😊 | 😊 | 😊 | 😊 | 😊 | 😊 | 😊 | 😊 | 😊 | 😊 |
| Liu 2016 | 😊 | 😞 | 😊 | 😊 | 😊 | 😊 | 😊 | 😞 | 😞 | 😞 | 😞 |
| Wang 2011 | 😞 | 😊 | 😊 | 😊 | 😊 | 😊 | 😊 | 😊 | 😊 | 😊 | 😊 |

**Table S6:** Subcomponent-specific results of the quality of reporting assessment in the reproducibility domain, specifically for the studies using RL. NA=not applicable

| Reference | Eligibility criteria | Treatment strategies | Outcome | Follow-up |  |  | Analysis plan |  |  |  |
| --- | --- | --- | --- | --- | --- | --- | --- | --- | --- | --- |
|  |  |  |  | Time zero | End of follow-up | Time resolution | Learning scheme | State space model | Environment model | Discount factor |
| Cheng 2019 | 😊 | 😊 | 😊 | 😊 | 😊 | 😊 | 😊 | 😊 | 😊 | 😊 |
| Eghbali 2021 | 😊 | 😊 | 😊 | 😊 | 😊 | 😊 | 😊 | 😊 | 😊 | 😊 |
| Jeter 2021 | 😊 | 😊 | 😊 | 😊 | 😊 | 😊 | 😊 | 😞 | 😊 | 😞 |
| Komorowski 2018 | 😊 | 😊 | 😊 | 😊 | 😊 | 😊 | 😊 | 😊 | 😊 | 😊 |
| Lejarza 2021 | 😊 | 😊 | 😊 | 😊 | 😊 | 😊 | 😊 | 😊 | 😊 | 😞 |
| Lin 2018 | 😊 | 😊 | 😊 | 😞 | 😞 | 😞 | 😊 | 😞 | 😊 | 😞 |
| Lopez-Martinez 2019 | 😊 | 😊 | 😊 | 😞 | 😞 | 😊 | 😊 | 😊 | 😊 | 😞 |
| Nemati 2016 | 😊 | 😊 | 😊 | 😊 | 😊 | 😊 | 😊 | 😊 | 😊 | 😞 |
| Padmanabhan 2015 | 😊 | 😊 | 😊 | NA | NA | NA | 😊 | 😊 | 😊 | 😞 |
| Padmanabhan 2017 | 😞 | 😊 | 😊 | NA | NA | NA | 😊 | 😊 | 😊 | 😞 |
| Peine 2021 | 😊 | 😊 | 😊 | 😊 | 😊 | 😊 | 😊 | 😊 | 😊 | 😊 |
| Petersen 2019 | 😊 | 😊 | 😊 | NA | NA | NA | 😊 | 😊 | 😊 | 😞 |
| Prasad 2017 | 😊 | 😊 | 😊 | 😊 | 😊 | 😊 | 😊 | 😊 | 😊 | 😞 |
| Raghu 2017 | 😊 | 😊 | 😊 | 😊 | 😊 | 😊 | 😊 | 😊 | 😊 | 😞 |
| Raghu 2018 | 😊 | 😊 | 😊 | 😊 | 😊 | 😊 | 😊 | 😞 | 😊 | 😞 |
| Roggeveen 2021 | 😊 | 😊 | 😊 | 😊 | 😊 | 😊 | 😊 | 😞 | 😊 | 😞 |
| Sinzinger 2005 | 😊 | 😊 | 😊 | NA | NA | NA | 😊 | 😊 | 😊 | 😊 |
| Sun 2021 | 😊 | 😊 | 😊 | 😊 | 😊 | 😊 | 😊 | 😞 | 😊 | 😊 |
| Weng 2017 | 😊 | 😊 | 😊 | 😊 | 😊 | 😊 | 😊 | 😊 | 😞 | 😊 |
| Zheng 2021 | 😊 | 😊 | 😊 | 😊 | 😊 | 😞 | 😊 | 😞 | 😊 | 😊 |

**Table S7:** reporting of assumptions assessment results per study. IPT=inverse probability of treatment

| Author | No unmeasured confounding |  |  | Positivity |  | Consistency |
| --- | --- | --- | --- | --- | --- | --- |
|  | Mentioned | Check for potential violations reported |  | Mentioned | Check for potential violations reported | Mentioned |
|  |  | Indirect method | Bias analysis |  | Examination of IPT weights distribution |  |
| Agodi 2017 | 😊 | 😞 | 😞 | 😞 | 😞 | 😞 |
| Althoff 2020 | 😊 | 😞 | 😊 | 😞 | 😞 | 😞 |
| Amer 2021 | 😊 | 😞 | 😞 | 😞 | 😞 | 😞 |
| Arabi 2018 | 😊 | 😊 | 😞 | 😞 | 😞 | 😞 |
| Arabi 2020 | 😊 | 😊 | 😞 | 😞 | 😞 | 😞 |
| Arnaud 2020 | 😊 | 😞 | 😞 | 😞 | 😊 | 😞 |
| Bailly 2015 | 😊 | 😊 | 😞 | 😊 | 😊 | 😊 |
| Bekaert 2011 | 😊 | 😞 | 😞 | 😞 | 😞 | 😞 |
| Chen 2021 | 😊 | 😊 | 😞 | 😞 | 😊 | 😞 |
| Cheng 2019 | 😞 | 😞 | 😞 | 😞 | 😞 | 😞 |
| De Bus 2020 | 😊 | 😊 | 😞 | 😊 | 😊 | 😞 |
| Delaney 2016 | 😊 | 😞 | 😞 | 😊 | 😊 | 😞 |
| Dupuis 2017 | 😊 | 😞 | 😊 | 😊 | 😊 | 😞 |
| Eghbali 2021 | 😊 | 😞 | 😞 | 😞 | 😞 | 😞 |
| Esperatti 2013 | 😞 | 😞 | 😞 | 😞 | 😞 | 😞 |
| Frencken 2018 | 😊 | 😞 | 😞 | 😞 | 😞 | 😞 |
| Jeter 2021 | 😞 | 😞 | 😞 | 😞 | 😞 | 😞 |
| Khanal 2012 | 😊 | 😞 | 😞 | 😊 | 😞 | 😞 |
| Klouwensberg 2014 | 😊 | 😊 | 😞 | 😞 | 😞 | 😞 |
| Komorowski 2018 | 😞 | 😞 | 😞 | 😞 | 😞 | 😞 |
| Lejarza 2021 | 😞 | 😞 | 😞 | 😞 | 😞 | 😞 |
| Li 2019 | 😞 | 😞 | 😞 | 😞 | 😊 | 😞 |
| Li 2020 | 😊 | 😞 | 😞 | 😞 | 😞 | 😞 |
| Libório 2020 | 😊 | 😞 | 😞 | 😊 | 😊 | 😞 |
| Lin 2018 | 😞 | 😞 | 😞 | 😞 | 😞 | 😞 |
| Liu 2016 | 😊 | 😞 | 😞 | 😞 | 😞 | 😊 |
| Lopez-Martinez 2019 | 😞 | 😞 | 😞 | 😞 | 😞 | 😞 |
| Mecklenburg 2021 | 😊 | 😞 | 😞 | 😞 | 😞 | 😞 |
| Muriel 2015 | 😊 | 😞 | 😞 | 😞 | 😞 | 😞 |
| Nemati 2016 | 😞 | 😞 | 😞 | 😞 | 😞 | 😞 |
| Ohbe 2018 | 😊 | 😊 | 😊 | 😞 | 😞 | 😞 |
| Ong 2015 | 😊 | 😞 | 😞 | 😞 | 😞 | 😞 |
| Ong 2016 | 😊 | 😊 | 😞 | 😞 | 😞 | 😞 |
| Padmanabhan 2015 | 😞 | 😞 | 😞 | 😞 | 😞 | 😞 |
| Padmanabhan 2017 | 😞 | 😞 | 😞 | 😞 | 😞 | 😞 |
| Parietti 2010 | 😞 | 😞 | 😞 | 😞 | 😞 | 😞 |
| Peine 2021 | 😊 | 😞 | 😞 | 😞 | 😞 | 😞 |
| Petersen 2019 | 😞 | 😞 | 😞 | 😞 | 😞 | 😞 |
| Pisani 2015 | 😊 | 😞 | 😞 | 😞 | 😞 | 😞 |
| Pouwels 2017 | 😊 | 😞 | 😞 | 😞 | 😊 | 😞 |
| Pouwels 2018 | 😊 | 😞 | 😞 | 😞 | 😊 | 😞 |
| Pouwels 2020 | 😊 | 😊 | 😞 | 😊 | 😊 | 😊 |
| Prasad 2017 | 😞 | 😞 | 😞 | 😞 | 😞 | 😞 |
| Raghu 2017 | 😊 | 😞 | 😞 | 😞 | 😞 | 😞 |
| Raghu 2018 | 😊 | 😞 | 😞 | 😞 | 😞 | 😞 |
| Roggeveen 2021 | 😞 | 😞 | 😞 | 😞 | 😞 | 😞 |
| Shahn 2020 | 😊 | 😊 | 😞 | 😊 | 😞 | 😊 |
| Shahn 2021 | 😊 | 😞 | 😞 | 😊 | 😊 | 😞 |
| Shelhamer 2021 | 😊 | 😊 | 😞 | 😞 | 😞 | 😞 |
| Sinzinger 2005 | 😞 | 😞 | 😞 | 😞 | 😞 | 😞 |
| Steen 2021 | 😊 | 😞 | 😞 | 😊 | 😊 | 😊 |
| Sun 2021 | 😞 | 😞 | 😞 | 😞 | 😞 | 😞 |
| Tacquard 2021 | 😞 | 😞 | 😞 | 😞 | 😞 | 😞 |
| Truche 2016 | 😊 | 😞 | 😊 | 😊 | 😊 | 😊 |
| Wang 2011 | 😊 | 😞 | 😞 | 😊 | 😞 | 😊 |
| Weng 2017 | 😞 | 😞 | 😞 | 😞 | 😞 | 😞 |
| Zhang, R. 2021 | 😞 | 😞 | 😞 | 😞 | 😊 | 😞 |
| Zhang, Z. 2018 | 😊 | 😞 | 😞 | 😞 | 😞 | 😞 |
| Zhang, Z. 2019 | 😊 | 😞 | 😞 | 😞 | 😊 | 😞 |
| Zheng 2021 | 😞 | 😞 | 😞 | 😞 | 😞 | 😞 |

## References

1 Komorowski M. Artificial intelligence in intensive care: are we there yet? Intensive Care Med 2019. https://link.springer.com/article/10.1007/s00134-019-05662-6.

2 Yoon JH, Pinsky MR, Clermont G. Artificial Intelligence in Critical Care Medicine. Crit Care 2022; 26. DOI:10.1186/s13054-022-03915-3.

3 Topol EJ. High-performance medicine: the convergence of human and artificial intelligence. Nat Med 2019; 25: 44–56.

4 van de Sande D, van Genderen ME, Huiskens J, Gommers D, van Bommel J. Moving from bytes to bedside: a systematic review on the use of artificial intelligence in the intensive care unit. Intensive Care Med 2021; 47: 750–60.

5 Komorowski M. Clinical management of sepsis can be improved by artificial intelligence: yes. Springer, 2020.

6 Hernán MA, Hsu J, Healy B. A Second Chance to Get Causal Inference Right: A Classification of Data Science Tasks. Chance 2019; 32: 42–9.

7 Fleuren LM, Klausch TLT, Zwager CL, et al. Machine learning for the prediction of sepsis: a systematic review and meta-analysis of diagnostic test accuracy. Intensive Care Med 2020; 46: 383–400.

8 Hernán M, Robins J. Causal Inference: What If. Boca Raton: Chapman & Hall/CRC., 2020.

9 Hernán MA, Robins JM. Using Big Data to Emulate a Target Trial When a Randomized Trial Is Not Available. Am J Epidemiol 2016; 183: 758–64.

10 Robins J, Hernan M. Estimation of the causal effects of time-varying exposures. In: Fitzmaurice G, Davidian M, Verbeke G, Molenberghs G, eds. Longitudinal Data Analysis. Chapman and Hall/CRC Press: New York, 2009: 553–99.

11 Mansournia MA, Etminan M, Danaei G, Kaufman JS, Collins G. Handling time varying confounding in observational research. BMJ 2017; 359: 1–6.

12 Rosenbaum PR, Rubin DB. The central role of the propensity score in observational studies for causal effects. Biometrika 1983; 70: 41–55.

13 Karim ME, Tremlett H, Zhu F, Petkau J, Kingwell E. Dealing With Treatment-Confounder Feedback and Sparse Follow-up in Longitudinal Studies: Application of a Marginal Structural Model in a Multiple Sclerosis Cohort. Am J Epidemiol 2021; 190: 908–17.

14 Daniel RM, Cousens SN, De Stavola BL, Kenward MG, Sterne JAC. Methods for dealing with time-dependent confounding. Stat Med 2013; 32: 1584–618.

15 Naimi AI, Cole SR, Kennedy EH. An introduction to g methods. Int J Epidemiol 2017; 46: 756–62.

16 Schisterman EF, Cole SR, Platf RW. Overadjustment bias and unnecessary adjustment in epidemiologic studies. Epidemiology 2009; 20: 488–95.

17 Greenland S. Quantifying Biases in Causal Models: Classical Confounding vs Collider-Stratification Bias. Epidemiology 2003; 14: 300–6.

18 Robins JM. A new approach to causal inference in mortality studies with a sustained exposure period—application to control of the healthy worker survivor effect. Math Model 1986; 7: 1393–512.

19 Sutton RS, Barto AG. Reinforcement learning: An introduction. MIT press, 2018.

20 Liu S, See KC, Ngiam KY, Celi LA, Sun X, Feng M. Reinforcement learning for clinical decision support in critical care: Comprehensive review. J Med Internet Res 2020; 22. DOI:10.2196/18477.

21 Page MJ, McKenzie JE, Bossuyt PM, et al. The PRISMA 2020 statement: an updated guideline for reporting systematic reviews. BMJ 2021; 372: n71.

22 Smit J., Krijthe JH, van Bommel J, Gommers DAMPJ, Reinders MJT, van Genderen ME. Answering ‘What If?’ in the intensive care unit: a protocol for a systematic review and critical appraisal of methodology. PROSPERO 2022 CRD42022324014. https://www.crd.york.ac.uk/prospero/display_record.php?ID=CRD42022324014.

23 von Elm E, Altman DG, Egger M, Pocock SJ, Gøtzsche PC, Vandenbroucke JP. Strengthening the Reporting of Observational Studies in Epidemiology (STROBE) statement: guidelines for reporting observational studies. BMJ 2007; 335: 806–8.

24 Xu S, Ross C, Raebel MA, Shetterly S, Blanchette C, Smith D. Use of stabilized inverse propensity scores as weights to directly estimate relative risk and its confidence intervals. Value Heal J Int Soc Pharmacoeconomics Outcomes Res 2010; 13: 273–7.

25 Cole SR, Hernán MA. Constructing inverse probability weights for marginal structural models. Am J Epidemiol 2008; 168: 656–64.

26 Gottesman O, Futoma J, Liu Y, et al. Interpretable Off-Policy Evaluation in Reinforcement Learning by Highlighting Influential Transitions. In: Proceedings of the 37th International Conference on Machine Learning. JMLR.org, 2020.

27 Lange T, Rod NH. Commentary: Causal models adjusting for time-varying confounding - please send more data. Int J Epidemiol 2019; 48: 265–7.

28 Van Der Weele TJ, Ding P. Sensitivity analysis in observational research: Introducing the E-Value. Ann Intern Med 2017; 167: 268–74.

29 Petersen ML, Porter KE, Gruber S, Wang Y, Van Der Laan MJ. Diagnosing and responding to violations in the positivity assumption. Stat Methods Med Res 2012; 21: 31–54.

30 Johnson AEW, Pollard TJ, Shen L, et al. MIMIC-III, a freely accessible critical care database. Sci data 2016; 3: 160035.

31 Shahn Z, Shapiro NI, Tyler PD, Talmor D, Lehman L-WH. Fluid-limiting treatment strategies among sepsis patients in the ICU: A retrospective causal analysis. Crit Care 2020; 24. DOI:10.1186/s13054-020-2767-0.

32 Shahn Z, Lehman L-WH, Mark RG, Talmor D, Bose S. Delaying initiation of diuretics in critically ill patients with recent vasopressor use and high positive fluid balance. Br J Anaesth 2021. DOI:10.1016/j.bja.2021.04.035.

33 Wang W, Scharfstein D, Wang C, Daniels M, Needham D, Brower R. Estimating the Causal Effect of Low Tidal Volume Ventilation on Survival in Patients with Acute Lung Injury. J R Stat Soc Ser C Appl Stat 2011; 60: 475–96.

34 Precup D, Sutton RS, Singh SP. Eligibility Traces for Off-Policy Policy Evaluation. ICML ‘00 Proc Seventeenth Int Conf Mach Learn 2000; : 759–66.

35 Hanna JP, Stone P, Niekum S. Bootstrapping with models: Confidence intervals for off-policy evaluation. Proc Int Jt Conf Auton Agents Multiagent Syst AAMAS 2017; 1: 538–46.

36 Le HM, Voloshin C, Yue Y. Batch policy learning under constraints. 36th Int Conf Mach Learn ICML 2019 2019; 2019-June: 6589–600.

37 Jiang N, Li L. Doubly robust off-policy value evaluation for reinforcement learning. 33rd Int Conf Mach Learn ICML 2016 2016; 2: 1022–35.

38 Gottesman O, Johansson F, Meier J, et al. Evaluating Reinforcement Learning Algorithms in Observational Health Settings. 2018; : 1–16.

39 Bekaert M, Timsit JF, Vansteelandt S, et al. Attributable mortality of ventilator-associated pneumonia: A reappraisal using causal analysis. Am J Respir Crit Care Med 2011; 184: 1133–9.

40 De Bus L, Depuydt P, Steen J, et al. Antimicrobial de-escalation in the critically ill patient and assessment of clinical cure: the DIANA study. Intensive Care Med 2020; 46: 1404–17.

41 Ong DSY, Spitoni C, Klein Klouwenberg Pmc, et al. Cytomegalovirus reactivation and mortality in patients with acute respiratory distress syndrome. Intensive Care Med 2016; 42: 333–41.

42 Pouwels KB, Vansteelandt S, Batra R, et al. Estimating the Effect of Healthcare-Associated Infections on Excess Length of Hospital Stay Using Inverse Probability-Weighted Survival Curves. Clin Infect Dis 2020; 71: E415–20.

43 Steen J, Vansteelandt S, de Bus L, et al. Attributable mortality of ventilator-associated pneumonia replicating findings, revisiting methods. Ann Am Thorac Soc 2021; 18: 830–7.

44 Li X, Klompas M, Menchaca JT, Young JG. Effects of daily treatment with acid suppressants for stress ulcer prophylaxis on risk of ventilator-Associated events. Infect Control Hosp Epidemiol 2019; 41: 187–93.

45 Komorowski M, Celi LA, Badawi O, Gordon AC, Faisal AA. The Artificial Intelligence Clinician learns optimal treatment strategies for sepsis in intensive care. Nat Med 2018; 24: 1716–20.

46 Peine A, Hallawa A, Bickenbach J, et al. Development and validation of a reinforcement learning algorithm to dynamically optimize mechanical ventilation in critical care. npj Digit Med 2021; 4. DOI:10.1038/s41746-021-00388-6.

47 Sinzinger ED, Moore B. Sedation of simulated ICU patients using reinforcement learning based control. Int J Artif Intell Tools 2005; 14: 137–56.

48 Althoff MD, Holguin F, Yang F, et al. Noninvasive Ventilation Use in Critically Ill Patients with Acute Asthma Exacerbations. Am J Respir Crit Care Med 2020; 202: 1520–30.

49 Dupuis C, de Montmollin E, Buetti N, et al. Impact of early corticosteroids on 60-day mortality in critically ill patients with COVID-19: A multicenter cohort study of the OUTCOMEREA network. PLoS One 2021; 16. DOI:10.1371/journal.pone.0255644.

50 Ohbe H, Jo T, Yamana H, Matsui H, Fushimi K, Yasunaga H. Early enteral nutrition for cardiogenic or obstructive shock requiring venoarterial extracorporeal membrane oxygenation: a nationwide inpatient database study. Intensive Care Med 2018; 44: 1258– 65.

51 Truche A-S, Darmon M, Bailly S, et al. Continuous renal replacement therapy versus intermittent hemodialysis in intensive care patients: impact on mortality and renal recovery. Intensive Care Med 2016; 42: 1408–17.

52 Bailly S, Bouadma L, Azoulay E, et al. Failure of empirical systemic antifungal therapy in mechanically ventilated critically ill patients. Am J Respir Crit Care Med 2015; 191: 1139– 46.

53 Sterne JA, Hernán MA, Reeves BC, et al. ROBINS-I: A tool for assessing risk of bias in non-randomised studies of interventions. BMJ 2016; 355: 1–7.

54 Watkins CJCH, Dayan P. Q-learning. Mach Learn 1992; 8: 279–92.

55 Nemati S, Ghassemi MM, Clifford GD. Optimal medication dosing from suboptimal clinical examples: a deep reinforcement learning approach. Annu Int Conf IEEE Eng Med Biol Soc IEEE Eng Med Biol Soc Annu Int Conf 2016; 2016: 2978–81.

56 Padmanabhan R, Meskin N. Reinforcement learning-based control for combined infusion of sedatives and analgesics. 2017 4th Int … 2017. https://ieeexplore.ieee.org/abstract/document/8102643/.

57 Padmanabhan R, Meskin N, Haddad WM. Closed-loop control of anesthesia and mean arterial pressure using reinforcement learning. Biomed Signal Process Control 2015; 22: 54–64.

58 Chakraborty B, Murphy S, Strecher V. Inference for non-regular parameters in optimal dynamic treatment regimes. Stat Methods Med Res 2010; 19: 317–43.

59 Arabi YM, Mandourah Y, Al-Hameed F, et al. Corticosteroid therapy for critically ill patients with middle east respiratory syndrome. Am J Respir Crit Care Med 2018; 197: 757–67.

60 Chakraborty B, Moodie EEM. Statistical Methods for Dynamic Treatment Regimes: Reinforcement Learning, Causal Inference, and Personalized Medicine. Springer New York, 2013 https://books.google.nl/books?id=p-u7BAAAQBAJ.

61 Li X, Young JG, Toh S. Estimating Effects of Dynamic Treatment Strategies in Pharmacoepidemiologic Studies with Time-Varying Confounding: a Primer. Curr Epidemiol Reports 2017; 4: 288–97.

62 Hernán MA, Lanoy E, Costagliola D, Robins JM. Comparison of dynamic treatment regimes via inverse probability weighting. Basic Clin Pharmacol Toxicol 2006; 98: 237–42.

63 Brower RG, Matthay MA, Morris A, Schoenfeld D, Thompson BT, Wheeler A. Ventilation with lower tidal volumes as compared with traditional tidal volumes for acute lung injury and the acute respiratory distress syndrome. N Engl J Med 2000; 342: 1301–8.

64 Tennant PWG, Murray EJ, Arnold KF, et al. Use of directed acyclic graphs (DAGs) to identify confounders in applied health research: review and recommendations. Int J Epidemiol 2021; 50: 620–32.

65 Greenland S, Pearl J, Robins JM. Causal diagrams for epidemiologic research. Epidemiology 1999; 10: 37–48.

66 Jackson JW. Diagnostics for confounding of time-varying and other joint exposures. Epidemiology 2016; 27: 859–69.

67 Gottesman O, Johansson F, Komorowski M, et al. Guidelines for reinforcement learning in healthcare. Nat Med 2019; 25: 16–8.

68 Sauer CM, Dam TA, Celi LA, et al. Systematic Review and Comparison of Publicly Available ICU Data Sets-A Decision Guide for Clinicians and Data Scientists. Crit Care Med 2022; 50: e581–8.

69 Precup D, Sutton RS, Singh SP. Eligibility Traces for Off-Policy Policy Evaluation. In: Proceedings of the Seventeenth International Conference on Machine Learning. San Francisco, CA, USA: Morgan Kaufmann Publishers Inc., 2000: 759–766.

70 Kaufman JS. Marginalia: Comparing adjusted effect measures. Epidemiology 2010; 21: 490–3.

71 Wallace MP, Moodie EEM. Doubly-robust dynamic treatment regimen estimation via weighted least squares. Biometrics 2015; 71: 636–44.

72 Wallace MP, Moodie EEM, Stephens DA. Dynamic treatment regimen estimation via regression-based techniques: Introducing R package reg. J Stat Softw 2017; 80. DOI:10.18637/jss.v080.i02.

73 Zhang Z, Zheng B, Liu N. Individualized fluid administration for critically ill patients with sepsis with an interpretable dynamic treatment regimen model. Sci Rep 2020; 10: 1–9.

74 Ma P, Liu J, Shen F, et al. Individualized resuscitation strategy for septic shock formalized by finite mixture modeling and dynamic treatment regimen. Crit Care 2021; 25: 1–16.

75 Clare PJ, Dobbins TA, Mattick RP. Causal models adjusting for time-varying confounding - A systematic review of the literature. Int J Epidemiol 2019; 48: 254–65.

76 Farmer RE, Kounali D, Walker AS, et al. Application of causal inference methods in the analyses of randomised controlled trials: A systematic review. Trials 2018; 19: 1–14.

77 Liu S, See KC, Ngiam KY, Celi LA, Sun X, Feng M. Reinforcement Learning for Clinical Decision Support in Critical Care: Comprehensive Review. J Med Internet Res 2020; 22: e18477.

78 Lederer DJ, Bell SC, Branson RD, et al. Control of confounding and reporting of results in causal inference studies. Ann Am Thorac Soc 2019; 16: 22–8.

79 Maslove DM, Leisman DE. Causal Inference From Observational Data: New Guidance From Pulmonary, Critical Care, and Sleep Journals. Crit Care Med 2019; 47: 1–2.

80 Self WH, Semler MW, Bellomo R, et al. Liberal Versus Restrictive Intravenous Fluid Therapy for Early Septic Shock: Rationale for a Randomized Trial. Ann Emerg Med 2018; 72: 457–66.

81 Hjortrup PB, Haase N, Bundgaard H, et al. Restricting volumes of resuscitation fluid in adults with septic shock after initial management: the CLASSIC randomised, parallel-group, multicentre feasibility trial. Intensive Care Med 2016; 42: 1695–705.

82 Khanal N, Marshall MR, Ma TM, Pridmore PJ, Williams AB, Rankin APN. Comparison of outcomes by modality for critically ill patients requiring renal replacement therapy: A single-centre cohort study adjusting for time-varying illness severity and modality exposure. Anaesth Intensive Care 2012; 40: 260–8.

## Appendix B: Reference list of all included studies

1 Agodi A, Barchitta M, Quattrocchi A, et al. Preventable proportion of intubation-associated pneumonia: Role of adherence to a care bundle. PLoS One 2017;12. doi:10.1371/journal.pone.0181170

2 Althoff MD, Holguin F, Yang F, et al. Noninvasive Ventilation Use in Critically Ill Patients with Acute Asthma Exacerbations. Am J Respir Crit Care Med 2020;202:1520–30. doi:10.1164/rccm.201910-2021OC

3 Amer M, Kamel AM, Bawazeer M, et al. Clinical characteristics and outcomes of critically ill mechanically ventilated COVID-19 patients receiving interleukin-6 receptor antagonists and corticosteroid therapy: a preliminary report from a multinational registry. Eur J Med Res 2021;26:1–12. doi:10.1186/s40001-021-00591-x

4 Arabi YM, Mandourah Y, Al-Hameed F, et al. Corticosteroid therapy for critically ill patients with middle east respiratory syndrome. Am J Respir Crit Care Med 2018;197:757–67. doi:10.1164/rccm.201706-1172OC

5 Arabi YM, Shalhoub S, Mandourah Y, et al. Ribavirin and Interferon Therapy for Critically Ill Patients with Middle East Respiratory Syndrome: A Multicenter Observational Study. Clin Infect Dis 2020;70:1837–44. doi:10.1093/cid/ciz544

6 Arnaud FCDS, Libório AB. Attributable nephrotoxicity of vancomycin in critically ill patients: A marginal structural model study. J Antimicrob Chemother 2020;75:1031–7. doi:10.1093/jac/dkz520

7 Bailly S, Bouadma L, Azoulay E, et al. Failure of empirical systemic antifungal therapy in mechanically ventilated critically ill patients. Am J Respir Crit Care Med 2015;191:1139–46. doi:10.1164/rccm.201409-1701OC

8 Bekaert M, Timsit JF, Vansteelandt S, et al. Attributable mortality of ventilator-associated pneumonia: A reappraisal using causal analysis. Am J Respir Crit Care Med 2011;184:1133–9. doi:10.1164/rccm.201105-0867OC

9 Chen H, Xie J, Su N, et al. Corticosteroid Therapy Is Associated With Improved Outcome in Critically Ill Patients With COVID-19 With Hyperinflammatory Phenotype. Chest 2021;159:1793–802. doi:10.1016/j.chest.2020.11.050

10 Cheng L-F, Prasad N, Engelhardt BE. An Optimal Policy for Patient Laboratory Tests in Intensive Care Units. Pac Symp Biocomput 2019;24:320–31.https://www.embase.com/search/results?subaction=viewrecord&id=L626748121&from=export

11 De Bus L, Depuydt P, Steen J, et al. Antimicrobial de-escalation in the critically ill patient and assessment of clinical cure: the DIANA study. Intensive Care Med 2020;46:1404–17. doi:10.1007/s00134-020-06111-5

12 Delaney JW, Pinto R, Long J, et al. The influence of corticosteroid treatment on the outcome of influenza A(H1N1pdm09)-related critical illness. Crit Care 2016;20. doi:10.1186/s13054-016-1230-8

13 Dupuis C, Garrouste-Orgeas M, Bailly S, et al. Effect of transfusion on mortality and other adverse events among critically ill septic patients: An observational study using a marginal structural Cox model. Crit Care Med 2017;45:1972–80. doi:10.1097/CCM.0000000000002688

14 Eghbali N, Alhanai T, Ghassemi MM. Patient-Specific Sedation Management via Deep Reinforcement Learning. Front Digit Heal 2021;3. doi:10.3389/fdgth.2021.608893

15 Esperatti M, Ferrer M, Giunta V, et al. Validation of Predictors of Adverse Outcomes in Hospital-Acquired Pneumonia in the ICU. Crit Care Med 2013;41:2151–61.

16 Frencken JF, Wittekamp BHJ, Plantinga NL, et al. Associations between enteral colonization with gram-negative bacteria and intensive care unit-acquired infections and colonization of the respiratory tract. Clin Infect Dis 2018;66:497–503. doi:10.1093/cid/cix824

17 Jeter R, Lehman L-W, Josef C, et al. Learning to Treat Hypotensive Episodes in Sepsis Patients Using a Counterfactual Reasoning Framework. medRxiv Published Online First: 2021. doi:10.1101/2021.03.03.21252863

18 Khanal N, Marshall MR, Ma TM, et al. Comparison of outcomes by modality for critically ill patients requiring renal replacement therapy: A single-centre cohort study adjusting for time-varying illness severity and modality exposure. Anaesth Intensive Care 2012;40:260–8. doi:10.1177/0310057×1204000208

19 Klouwenberg PMCK, Zaal IJ, Spitoni C, et al. The attributable mortality of delirium in critically ill patients: Prospective cohort study. BMJ 2014;349:1–10. doi:10.1136/bmj.g6652

20 Komorowski M, Celi LA, Badawi O, et al. The Artificial Intelligence Clinician learns optimal treatment strategies for sepsis in intensive care. Nat Med 2018;24:1716–20. doi:10.1038/s41591-018-0213-5

21 Lejarza F, Calvert J, Attwood MM, et al. Optimal discharge of patients from intensive care via a data-driven policy learning framework. arXiv Prepr arXiv … Published Online First: 2021.https://arxiv.org/abs/2112.09315

22 Li X, Klompas M, Menchaca JT, et al. Effects of daily treatment with acid suppressants for stress ulcer prophylaxis on risk of ventilator-Associated events. Infect Control Hosp Epidemiol 2019;41:187–93. doi:10.1017/ice.2019.323

23 Li Y, Meng Q, Rao X, et al. Corticosteroid therapy in critically ill patients with COVID-19: a multicenter, retrospective study. Crit Care 2020;24. doi:10.1186/s13054-020-03429-w

24 Liborio AB, Barbosa ML, Sa VB, et al. Impact of loop diuretics on critically ill patients with a positive fluid balance. Anaesthesia 2020;75 Suppl 1:e134–42. doi:10.1111/anae.14908 LB - 31903562

25 Lin R, Stanley MD, Ghassemi MM, et al. A Deep Deterministic Policy Gradient Approach to Medication Dosing and Surveillance in the ICU. Conf Proc. Annu Int Conf IEEE Eng Med Biol Soc IEEE Eng Med Biol Soc Annu Conf 2018;2018:4927–31. doi:10.1109/EMBC.2018.8513203

26 Liu Q, Henry KE, Xu Y, et al. Using Causal Inference to Estimate What-if Outcomes for Targeting Treatments. 2016.

27 Lopez-Martinez D, Eschenfeldt P, Ostvar S, et al. Deep Reinforcement Learning for Optimal Critical Care Pain Management with Morphine using Dueling Double-Deep Q Networks. Conf Proc. Annu Int Conf IEEE Eng Med Biol Soc IEEE Eng Med Biol Soc Annu Conf 2019;2019:3960–3. doi:10.1109/EMBC.2019.8857295

28 Mecklenburg A, Stamm J, Angriman F, et al. Impact of therapeutic hypothermia on bleeding events in adult patients treated with extracorporeal life support peri-cardiac arrest. J Crit Care 2021;62:12–8. doi:10.1016/j.jcrc.2020.11.008

29 Muriel A, Peñuelas O, Frutos-Vivar F, et al. Impact of sedation and analgesia during noninvasive positive pressure ventilation on outcome: a marginal structural model causal analysis. Intensive Care Med 2015;41:1586–600. doi:10.1007/s00134-015-3854-6

30 Nemati S, Ghassemi MM, Clifford GD. Optimal medication dosing from suboptimal clinical examples: a deep reinforcement learning approach. Conf Proc. Annu Int Conf IEEE Eng Med Biol Soc IEEE Eng Med Biol Soc Annu Conf 2016;2016:2978–81. doi:10.1109/EMBC.2016.7591355

31 Ohbe H, Jo T, Yamana H, et al. Early enteral nutrition for cardiogenic or obstructive shock requiring venoarterial extracorporeal membrane oxygenation: a nationwide inpatient database study. Intensive Care Med 2018;44:1258–65. doi:10.1007/s00134-018-5319-1

32 Ong DSY, Bonten MJM, Safdari K, et al. Epidemiology, management, and risk-adjusted mortality of ICU-acquired enterococcal bacteremia. Clin Infect Dis 2015;61:1413–20. doi:10.1093/cid/civ560

33 Ong DSY, Spitoni C, Klein Klouwenberg Pmc, et al. Cytomegalovirus reactivation and mortality in patients with acute respiratory distress syndrome. Intensive Care Med 2016;42:333–41. doi:10.1007/s00134-015-4071-z

34 Padmanabhan R, Meskin N, Haddad WM. Closed-loop control of anesthesia and mean arterial pressure using reinforcement learning. Biomed Signal Process Control 2015;22:54–64. doi:10.1016/j.bspc.2015.05.013

35 Padmanabhan R, Meskin N. Reinforcement learning-based control for combined infusion of sedatives and analgesics. 2017 4th Int … Published Online First: 2017.https://ieeexplore.ieee.org/abstract/document/8102643/

36 Parienti JJ, Dugué AE, Daurel C, et al. Continuous renal replacement therapy may increase the risk of catheter infection. Clin J Am Soc Nephrol 2010;5:1489–96. doi:10.2215/CJN.02130310

37 Peine A, Hallawa A, Bickenbach J, et al. Development and validation of a reinforcement learning algorithm to dynamically optimize mechanical ventilation in critical care. npj Digit Med 2021;4. doi:10.1038/s41746-021-00388-6

38 Petersen BK, Yang J, Grathwohl WS, et al. Deep Reinforcement Learning and Simulation as a Path Toward Precision Medicine. J Comput Biol a J Comput Mol cell Biol 2019;26:597–604. doi:10.1089/cmb.2018.0168

39 Pisani MA, Araujo KLB, Murphy TE. Association of cumulative dose of haloperidol with next-day delirium in older medical ICU patients. Crit Care Med 2015;43:996–1002. doi:10.1097/CCM.0000000000000863

40 Pouwels KB, Van Kleef E, Vansteelandt S, et al. Does appropriate empiric antibiotic therapy modify intensive care unit-acquired Enterobacteriaceae bacteraemia mortality and discharge? J Hosp Infect 2017;96:23–8. doi:10.1016/j.jhin.2017.03.016

41 Pouwels KB, Vansteelandt S, Batra R, et al. Intensive care unit (ICU)-acquired bacteraemia and ICU mortality and discharge: addressing time-varying confounding using appropriate methodology. J Hosp Infect 2018;99:42–7. doi:10.1016/j.jhin.2017.11.011

42 Pouwels KB, Vansteelandt S, Batra R, et al. Estimating the Effect of Healthcare-Associated Infections on Excess Length of Hospital Stay Using Inverse Probability-Weighted Survival Curves. Clin Infect Dis 2020;71:E415–20. doi:10.1093/cid/ciaa136

43 Prasad N, Cheng LF, Chivers C, et al. A Reinforcement Learning Approach to Weaning of Mechanical Ventilation in Intensive Care Units. ArXiv170406300 Cs. 2017 Apr 20. 2019.

44 Raghu A, Komorowski M, Ahmed I, et al. Deep Reinforcement Learning for Sepsis Treatment. arXiv Prepr arXiv181109602 Published Online First: 2018.https://arxiv.org/abs/1811.09602

45 Raghu A, Komorowski M, Singh S. Model-Based Reinforcement Learning for Sepsis Treatment. Published Online First: 2018.http://arxiv.org/abs/1811.09602

46 Roggeveen L, el Hassouni A, Ahrendt J, et al. Transatlantic transferability of a new reinforcement learning model for optimizing haemodynamic treatment for critically ill patients with sepsis. Artif Intell Med 2021;112. doi:10.1016/j.artmed.2020.102003

47 Shahn Z, Shapiro NI, Tyler PD, et al. Fluid-limiting treatment strategies among sepsis patients in the ICU: A retrospective causal analysis. Crit Care 2020;24. doi:10.1186/s13054-020-2767-0

48 Shahn Z, Lehman L-WH, Mark RG, et al. Delaying initiation of diuretics in critically ill patients with recent vasopressor use and high positive fluid balance. Br J Anaesth Published Online First: 2021. doi:10.1016/j.bja.2021.04.035

49 Shelhamer MC, Wesson PD, Solari IL, et al. Prone Positioning in Moderate to Severe Acute Respiratory Distress Syndrome Due to COVID-19: A Cohort Study and Analysis of Physiology. J Intensive Care Med 2021;36:241–52. doi:10.1177/0885066620980399

50 Sinzinger ED, Moore B. Sedation of simulated ICU patients using reinforcement learning based control. Int J Artif Intell Tools 2005;14:137–56.

51 Steen J, Vansteelandt S, de Bus L, et al. Attributable mortality of ventilator-associated pneumonia replicating findings, revisiting methods. Ann Am Thorac Soc 2021;18:830–7. doi:10.1513/AnnalsATS.202004-385OC

52 Sun C, Hong S, Song M, et al. Personalized vital signs control based on continuous action-space reinforcement learning with supervised experience. Biomed Signal Process Control 2021;69:102847. doi:10.1016/j.bspc.2021.102847

53 Tacquard C, Mansour A, Godon A, et al. Impact of High-Dose Prophylactic Anticoagulation in Critically Ill Patients With COVID-19 Pneumonia. Chest 2021;159:2417–27. doi:10.1016/j.chest.2021.01.017

54 Truche A-S, Darmon M, Bailly S, et al. Continuous renal replacement therapy versus intermittent hemodialysis in intensive care patients: impact on mortality and renal recovery. Intensive Care Med 2016;42:1408–17. doi:10.1007/s00134-016-4404-6

55 Wang W, Scharfstein D, Wang C, et al. Estimating the Causal Effect of Low Tidal Volume Ventilation on Survival in Patients with Acute Lung Injury. J R Stat Soc Ser C Appl Stat 2011;60:475–96. doi:10.1111/j.1467-9876.2010.00757.x

56 Weng WH, Gao M, He Z, et al. Representation and reinforcement learning for personalized glycemic control in septic patients. arXiv Prepr arXiv … Published Online First: 2017.https://arxiv.org/abs/1712.00654

57 Zhang R, Chen H, Gao Z, et al. The Effect of Loop Diuretics on 28-Day Mortality in Patients With Acute Respiratory Distress Syndrome. Front Med 2021;8. doi:10.3389/fmed.2021.740675

58 Zhang Z, Zhu C, Mo L, et al. Effectiveness of sodium bicarbonate infusion on mortality in septic patients with metabolic acidosis. Intensive Care Med 2018;44:1888–95. doi:10.1007/s00134-018-5379-2

59 Zhang Z, Mo L, Ho KM, et al. Association between the use of sodium bicarbonate and mortality in acute kidney injury using marginal structural COx model. Crit Care Med 2019;47:1402–8. doi:10.1097/CCM.0000000000003927

60 Zheng H, Zhu J, Xie W, et al. Reinforcement learning assisted oxygen therapy for COVID-19 patients under intensive care. BMC Med Inform Decis Mak 2021;21:350. doi:10.1186/s12911-021-01712-6

